# Hypothesis-free detection of gene-interaction effects on biomarker concentration in UK Biobank using variance prioritisation

**DOI:** 10.1101/2022.01.05.21268406

**Authors:** Matthew S. Lyon, Louise A. C. Millard, George Davey Smith, Tom R. Gaunt, Kate Tilling

## Abstract

Blood biomarkers include disease intervention targets that may interact with genetic and environmental factors resulting in subgroups of individuals who respond differently to treatment. Such interactions may be observed in genetic effects on trait variance. Variance prioritisation is an approach to identify genetic loci with interaction effects by estimating their association with trait variance, even where the modifier is unknown or unmeasured. Here, we develop and evaluate a regression-based Brown-Forsythe test and variance effect estimate to detect such interactions. We provide scalable open-source software (varGWAS) for genome-wide association analysis of SNP-variance effects (https://github.com/MRCIEU/varGWAS) and apply our software to 30 blood biomarkers in UK Biobank. We find 468 variance quantitative trait loci across 24 biomarkers and follow up findings to detect 82 gene-environment and six gene-gene interactions independent of strong scale or phantom effects. Our results replicate existing findings and identify novel epistatic effects of *TREH* rs12225548 x *FUT2* rs281379 and *TREH* rs12225548 x *ABO* rs635634 on alkaline phosphatase and *ZNF827* rs4835265 x *NEDD4L* rs4503880 on gamma glutamyltransferase. These data could be used to discover possible subgroup effects for a given biomarker during preclinical drug development.

## Introduction

Blood biomarkers provide valuable information for diagnosis and prognosis of disease^1^, insight into biological mechanisms^2^, and a source of causal modifiable risk factors which may be intervened upon to create therapies^1^. For example, lipids, glucose, and urate have become successful therapeutic targets for cardiovascular disease^3^, type 2 diabetes^4^, and gout^5^, respectively, among others. However, as biomarkers are complex traits they are affected by genetic and environmental factors which may interact producing gene-gene (GxG, epistasis) or gene-environment (GxE) effects^6^. Intervening on biomarkers which have an interaction effect on disease outcome will produce subgroup effects with individual variation in response to treatment dependent on the modifier^7^. Identifying these interactions may contribute to stratified medicine which aims to provide optimum treatments and preventative advice for disease based on individual characteristics^6^.

Detecting interaction effects can be challenging. Statistical power to detect an interaction is lower than for main effects; for randomised control trials the sample size needed to detect an interaction with equal sized subgroups is around four times the size needed to detect the main effect of the same magnitude^7,8^. Low power is exacerbated by multiple testing correction that is essential to account for evaluating the large numbers of candidate modifiers. To reduce multiple testing, pairwise interaction analyses of SNPs with moderate main effects can be performed. However, this approach could miss subgroups with an effect in only one group or opposing effect directionality (known as qualitative interaction effects^7^) hence weaker overall effects, yet these offer the most potential for stratified medicine. An alternative approach to select SNPs for GxG/GxE testing is variance prioritisation^9,10^ which identifies differences in outcome variance across genotype levels (variance quantitative trait loci, vQTL). Although not conclusive evidence, this observation is consistent with a SNP-interaction effect^11^ and detection of vQTLs does not require the modifier to be measured^11^.

Variance QTLs can arise as a consequence of heterogeneous mean effects that could occur from changing environment, background genetics and temporal regulation^11^. Among the first reported vQTL effects in humans was rs7202116 (*FTO* locus), associated with a large change in variance (as well as mean) of body mass index (BMI)^12^. More recently, systematic testing of vQTL effects on 13 quantitative traits in UK Biobank and subsequent GxE testing identified 16 GxE effects modified by age, sex, physical activity, sedentary behaviour, and smoking^13^. Variance QTLs have also been identified for gene expression^14^, DNA methylation^15^, Vitamin D^16^ and facial morphology^17^. To date, gene-interaction studies have mostly focused on testing a small number of candidate interactions, but hypothesis-free testing of vQTL effects on blood biomarkers could lead to the identification of unanticipated intervention targets with subgroup effects.

Existing studies of vQTLs have employed a range of methods^13,18–20^. Wang *et al* compared the power and type I error of four widely used variance tests and found the median variant of Levene’s test^21^ also known as the Brown-Forsythe test^13,22^ to be most robust. However, this test does not allow for inclusion of covariates or continuous genotype data (i.e., imputed allelic dose) and does not provide an effect estimate, all of which are limitations when applied in a GWAS. However, the Brown-Forsythe test can be reformulated using least-absolute deviation^15,23^ (LAD) regression using the same structure as the Glejser test^24^. Regression-based variance tests offer greater flexibility to overcome these limitations. Recent developments in LAD regression have vastly reduced the computational burden for large high-dimensional datasets^25^.

In this study we compare the utility of the original Brown-Forsythe test and our LAD regression-based reformulation of the Brown-Forsythe test (LAD-BF) to detect SNP-interaction effects under simulation and develop scalable open-source software (https://github.com/MRCIEU/varGWAS) for performing variance GWAS using the latter. We apply our regression-based model to estimate SNP effects on the variance of 30 blood biomarkers in∼337K UK Biobank participants and follow up vQTLs with formal interaction tests to detect GxG and GxE interactions.

## Material and methods

### Original Brown-Forsythe test

The Brown-Forsythe^22^ test (median variant of Levene’s test^21^) refers to the original published non-parametric test and will be used throughout. We applied the Brown-Forsythe test to detect differences in trait variability across the three genotypic groups.

The test statistic *W* is F-distributed *F*(2, *N* −3) given by:

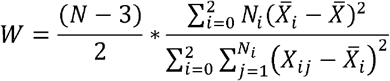

Where *N* is the total number of observations. *N*_*i*_ is the number of observations with the *i*th genotype group {0, 1, 2}. *X*_*ij*_ is the absolute residual of the outcome for the *j*th observation in the *i*th genotype group from the median.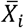 is the mean of *X*_*ij*_ for the *i*th genotype group and 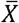 is the mean of *X*_*ij*_ across genotype groups.

All analyses of the original Brown-Forsythe test used the omic-data-based complex trait analysis (OSCA) software package^13,26^ which additionally produces a variance effect estimate derived from the test P-value assuming linearity between the SNP and outcome variance^27^.

### LAD-BF test

Our reformulated regression-based Brown-Forsythe test uses LAD regression of outcome *Y* on independent variable *X* to estimate the residuals adjusting for any covariates:

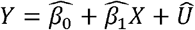

Where *X* is the genotype measured by continuous (expected value from genotype imputation) or ordinal (directly genotyped) variable and *Û* is the residual of this first-stage model.

A second-stage ordinary least squares (OLS) model regressed the absolute residuals | *Û* | of the first-stage model on the genotype values coded as dummy variables (genotype expected values were rounded to the nearest whole number resulting in some loss of precision) including any covariates given in the first-stage model:

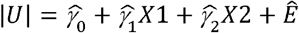

The test P-value was estimated from an F-test comparing the second-stage residual sum of squares to an intercept-only model to test the null hypothesis of variability homogeneity across genotypes.

SNP effects on trait variance were calculated from second-stage regression coefficients which are estimates of mean-absolute deviation. This transformation assumes trait normality.

The *var* (*Y*|*G* == 1) was estimated using:

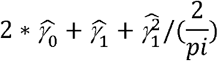

The *var* (*Y*|*G* == 2) was estimated using:

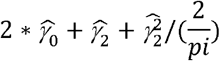

The standard error of the variance effect was estimated using the delta method^28^ and heteroscedastic-consistent standard errors for the second-stage model coefficients^29^.

LAD regression was implemented using the majorise-minimisation^25,30^ (MM) model with default values for iterations (200) and tolerance (0.001) and first-stage OLS regression coefficients provided as initial values.

### Software

The LAD-BF test was implemented in varGWAS available in C++ v1.2.3 and R v1.0.0 (refer to the code and data availability section). The MM model used functionality from the cqrReg R-package^25^ (https://cran.r-project.org/web/packages/cqrReg/index.html). OLS and general matrix functionality were provided with Eigen v3.4.0^31^. BGEN file processing used the BGEN library^32^ v1.1.6.

The original Brown-Forsythe test used the OSCA software package v0.46^13,26^. Simulations and follow up UK Biobank analyses were performed using R v3.6.0.

### Simulations

The bias and statistical power of the two Brown-Forsythe tests were evaluated through a series of simulation studies reported using the ADEMP structure^33^ (**Table 1 & Supplemental Material and Methods**).

**Table 1.** Simulation studies of the Brown-Forsythe test.

| **Figure** | **Aim** | **Conditions** |
| --- | --- | --- |
| **1** | Determine variance effect estimate accuracy and confidence interval coverage | Increasing interaction effect size. |
| **S2** | Estimate power of variance tests to detect interaction effects using normal/non-normal outcomes | Increasing interaction effect size.<br>Normal, log-normal, and t-distribution residuals |
| **S3** | Estimate type I error of variance tests with normal/non-normal outcomes | Increasing SNP minor allele frequency.<br>Normal, log-normal, t-distribution, and mixed-normal residuals |
| **S4** | Adjusting for interaction effect on variance effect estimate | With/without adjustment for modifier and interaction term in first-stage model |
| **S5** | Estimate model runtime performance | Increasing number of CPU threads |

SNP, single-nucleotide polymorphism. Log, natural logarithm. CPU, central processing unit.

### Participants

UK Biobank is a large prospective cohort study of approximately 500,000 UK participants aged 37-73 at recruitment^34^. Recruitment took place between 2006-2010 from across the UK. Measures were collected on lifestyle, socio-demographics, physical parameters, health-related factors, and biological samples for genetic testing and biomarker measurements. Ethical approval for the UK Biobank study was granted by the National Research Ethics Service (NRES) Committee North West (ref 11/NW/0382). All analyses were performed under approved UK Biobank project 15825 (dataset ID 33352).

### Genetic data

Genetic array data were available on 488,377 participants measured using a combination of UK Biobank Axiom™ array (n=438,398) and UK BiLEVE array (n=49,979). Genotype imputation was performed using a reference set combined with UK10K haplotypes and HRC reference panels with the IMPUTE2^35^ software as described^36^. The following SNPs were removed from analysis leaving a total of 6,812,700: multi-allelic loci, minor allele frequency < 5%, Hardy-Weinberg violations (P < 1 × 10^−5^), genotype missing rate >5%, low imputation score (INFO < 0.3) and *HLA* locus (hg19/GRCh37 chr6:23477797-38448354).

### Quality control

We applied standard exclusion criteria (**Figure S1**) to remove genotype-phenotype sex mismatches, aneuploidies, and outliers for missingness or heterozygosity as previously described^36^ leaving n=486,565 participants. To ensure data independence, closely related subjects were removed as described elsewhere^36^ leaving n=407,176 participants. Finally, ‘non-white British’ participants defined using published methodology^36^ were removed to avoid confounding by population stratification providing a total sample size of n=377,076.

### Phenotypes

UK Biobank measures of 30 serum biochemistry markers were available for approximately 500k participants. Each measure was chosen based on being an established risk factor for disease, a clinical diagnostic measure or because it characterises a phenotype that is not well assessed by other approaches as described in the UK Biobank documentation^37^. Quantification and quality control was performed as previously described^37^. Total physical activity was calculated by summing self-reported duration of walking, moderate and vigorous activity collected using the International Physical Activity Questionnaire as described^38^. For each analysis participants with missing data were removed. All continuous outcomes were SD normalised.

### Genome-wide association studies (GWAS)

GWAS of biomarker variability were performed using our LAD regression-based Brown-Forsythe test adjusted for age, sex, and the top ten genetic principal components in first and second regression models. We removed outlier biomarker values with a Z-score > 5SD from the mean to control type I error inflation as previously described^13^. Quality control was undertaken visually using Q-Q plots to check for a departure of P-value distribution from that expected under the null. Independent vQTLs were identified by clumping GWAS loci that passed the experiment-wise genome-wide evidence threshold P < 1.67 × 10^−9^ (Bonferroni correction of standard GWAS threshold: p = 5 × 10^−8^ / 30) using the OpenGWAS API^39^ with default R^2^ threshold of 0.001 and 1000 genomes phase 3 European ancestry^40^.

### Gene interaction test

Independent vQTLs (see above) were tested for interaction effects on additive and multiplicative scales using heteroscedasticity-consistent standard errors^29^ adjusted for age, sex, and top ten genetic principal components. To ensure effects were robust to phantom effects^41^, we performed sensitivity analyses adjusting for fine-mapped main effects identified using SuSiE^42^ (**Supplemental Material and Methods**). Interactions surpassing genome-wide association significance (P < 5 × 10^−8^) on additive and multiplicative scales that did not strongly attenuate with adjustment for fine-mapped main effects were prioritised for subgroup analyses. GxG effects were identified through interaction testing with independent (R^2^ < 0.001) vQTLs excluding pairwise combinations of vQTLs within a 10Mb window as previously described^13^. GxE testing was performed using candidate modifiers: age, sex, body mass index, alcohol intake, smoking status, physical activity, daily sugar intake, and daily fat intake.

### Subgroup analyses

Subgroup effects of top interaction effects were presented by estimating the SNP effect on the outcome stratified by modifier using heteroscedasticity-consistent standard errors^29^ adjusted for age, sex and top ten genetic principal components. Modifiers were rounded genetic dosage values or prepared by dichotomisation as follows: below or above the median value for continuous variables (group [G] 1, below median; G2, median or greater), ever (G1) vs never (G2) smoker, alcohol intake once a week or more (G1) vs less than once a week on average (G2), males (G1) vs females (G2). Subgroup effects are presented along with the SNP-variance estimates adjusted for age, sex and top ten genetic principial components with and without adjustment for the interaction term (variance effects were not adjusted for sex when sex was the modifier).

### Gene annotation

Variance QTLs were annotated with the nearest gene using the closest function of bedtools^43^ (v2.3.0) and Ensembl v104 (GRCh37) protein-coding features which were filtered to retain HUGO^44^ valid identifiers. The following annotations were recoded based on expression QTL evidence^45,46^: rs4530622 *SLC2A9*, rs11244061 *ABO*, rs71633359 *HSD17B13*, rs28413939 *TREH*, rs281379 *FUT2*, rs635634 *ABO*, rs964184 *APOA5*.

## Results

### Simulated power and type I error to detect interaction effects by change in variance

The power to detect a difference in trait variability due to an interaction effect was low and equivalent for both methods (**Figure S2**). Suppose a SNP has a main effect on a normally distributed outcome detectable with 80% power, then 10x the sample size needed to detect the main effect was required to detect the interaction with only 50% power assuming the interaction was half the size of the main effect. Positive skew and kurtosis reduced power. Both methods had equally well controlled type I error (**Figure S3**).

### Simulated variance effect estimate and confidence interval coverage

Under a simulated linear effect of genotype on outcome variance both methods gave the correct effect estimate and 95% confidence interval coverage (**Figure 1**). However, when the difference in variance was a consequence of an interaction effect, the relationship between the genotype and outcome variance was non-linear and dependent on the modifier. Under these conditions, the variance effect estimate produced using OSCA^26,27^ from the Brown-Forsythe test P-value gave the incorrect effect size while LAD-BF produced the correct estimate albeit with slightly elevated coverage.

**Figure 1.**
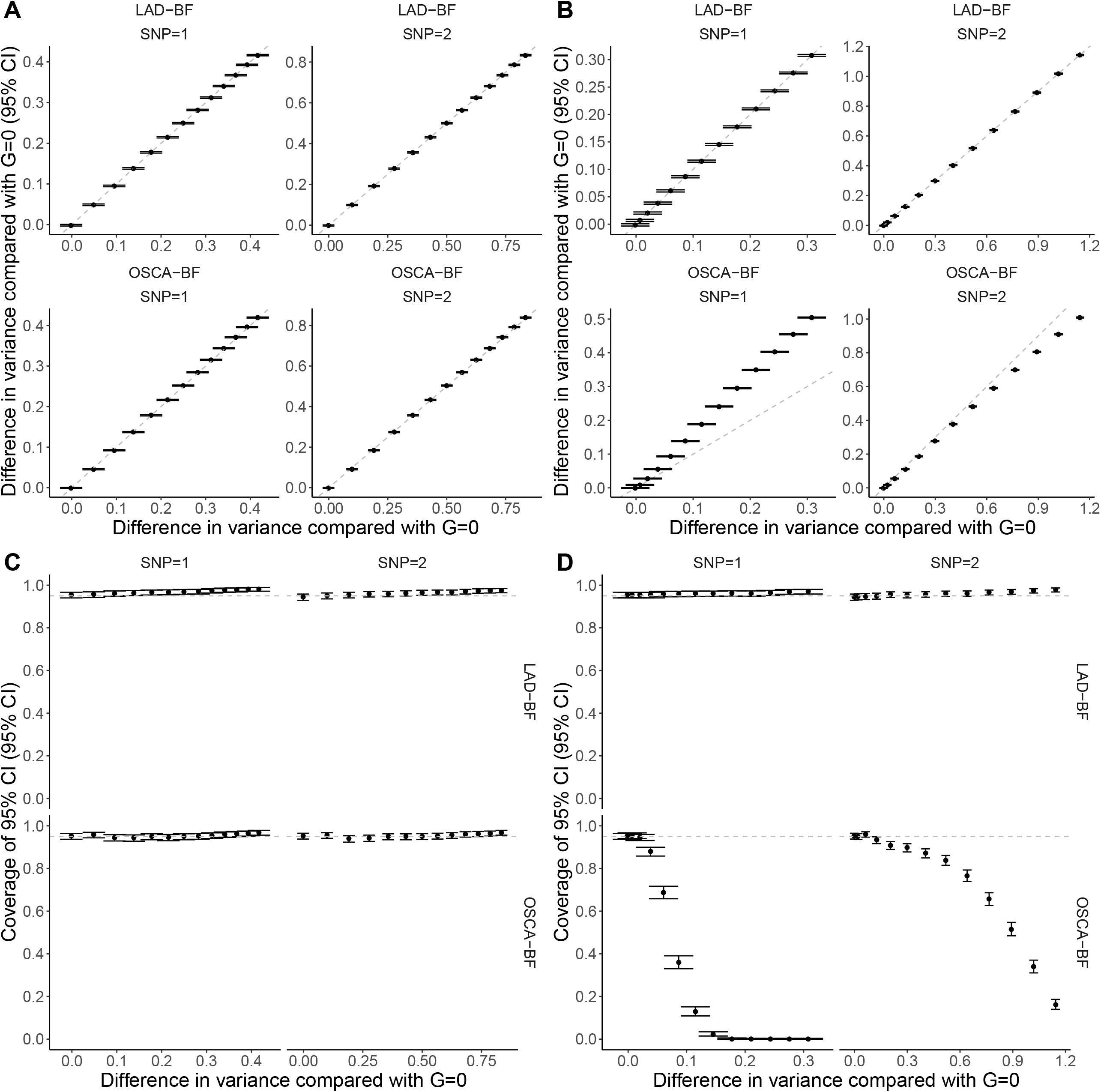
Variance effect estimate accuracy and confidence interval coverage. Variance effect estimate accuracy (A, B) and 95% confidence interval coverage (C, D) of simulated genotypes with linear effect on outcome variance (A, C) or interaction effect (B, D). LAD-BF, least-absolute deviation regression Brown-Forsythe. OSCA-BF, original Brown-Forsythe test implemented in OSCA^26^ including effect estimate derived from the test P-value^27^. CI, confidence interval.

### Adjusting the LAD-BF test for an interaction effect through simulation

We simulated an interaction effect and compared the LAD-BF test P-value distributions with and without adjusting for the simulated interaction (**Figure S4**). Including the interaction term in the first-stage regression model completely attenuated the variance test statistic. After identifying an interaction at a variance locus this approach could be applied to determine if additional strong interaction effects exist and could be used in a stepwise regression fashion until all interaction effects are identified.

### Runtime performance

Increasing the number of CPU threads reduced the total runtime of both methods to process 1000 SNPs (**Figure S5**). For the C++ implementation of LAD-BF in varGWAS, the lowest average runtime was 13.6 second (95% CI 13.5, 13.7) using four threads of an Intel(R) Xeon(R) CPU E5-2680 v4 @ 2.40GHz. Under the same conditions, the original Brown-Forsythe test implemented in OSCA was 1.78x faster (7.61 seconds [95% CI 7.60, 7.63]).

### GWAS of variance effects in UK Biobank

We identified 468 independent (R^2^ < 0.001) vQTLs influencing 24 biomarkers (**Figure S6**,**Figure S7 & Table S1**) using an experiment-wise P-value threshold of 1.67 × 10^−9^ (5 × 10^−8^ / 30) and no variance effects for albumin, calcium, oestradiol, phosphate, rheumatoid factor, or total protein. Oestradiol and rheumatoid factor were measured on a subset of n=76,674 and n=41,315 participants respectively and therefore were less well powered to detect effects. Of these vQTLs, 270 (57.7%) had suggestive evidence for a variance effect on the log scale (P < 5 × 10^−5^) and 453 (96.8%) had a mean effect (P < 5 × 10^−8^). The low concordance between natural and log scales and high concordance between mean and variance effects suggests the presence of mean-variance relationships which is a likely consequence of extreme non-normality for some of the trait distributions (**Figure S8**).

### Gene-environment interaction effects (GxE)

We detected 139 additive and 104 multiplicative GxE effects (P < 5 × 10^−8^; **Figure S9 & Figure S10**). Adjusting the additive effects for fine-mapped main effects (**Figure S11**) led to a small increase in *UGT1A8* rs2741047 x sex on direct bilirubin to 0.037 SD (95% CI 0.032, 0.042) from 0.028 SD (95% CI 0.023, 0.033) and minor attenuation of *MAP3K4* rs1247295 x sex on lipoprotein a to -0.011 SD (95% CI -0.015, -0.007) from -0.016 SD (95% CI -0.021, -0.010). These findings could reflect the presence of large main effects in imperfect linkage disequilibrium with the index SNP which is known to inflate/deflate test statistics^41^.

We prioritised 82 GxE effects with evidence on both scales (P < 5 × 10^−8^) to avoid spurious interactions dependent on scale (**Table S2**). Of these BMI (n=35), sex (n=27) and age (n=17) modified most effects and smoking status (n=2) and alcohol intake (n=1) fewer. We also tested for interaction by physical activity, and sugar and fat intake but identified little evidence of interactions. The largest effects (**Figure 2**) were: *PNPLA3* rs738409 x BMI on alanine aminotransferase (ALT; 0.08 SD [95% CI 0.08, 0.09]), *SLC2A9* rs938555 x sex on urate (−0.08 SD [95% CI -0.09, -0.08]), *APOE* rs1065853 x sex on low-density lipoprotein (LDL; 0.06 SD [95% CI 0.05, 0.07]), *SHBG* rs1799941 x sex on testosterone (0.06 SD [95% CI 0.06, 0.06]) and *TM6SF2* rs58542926 x BMI on ALT (0.05 SD [95% CI 0.04, 0.06]). Adjusting the variance effect for the interaction term (**Figure 2**) led to attenuation of *PNPLA3* rs738409 and *TM6SF2* rs58542926 on ALT and *SHBG* rs1799941 on testosterone but strong variance effects on ALT remained at *PNPLA3* rs738409 (LAD-BF P_adjust = 1.0 × 10^−73^) and *TM6SF2* rs58542926 (LAD-BF P_adjust = 1.84 × 10^−8^). There was no strong variance attenuation of *APOE* rs1065853 on LDL or *SLC2A9* rs938555 on urate following adjustment for the interaction (**Figure 2**).

**Figure 2.**
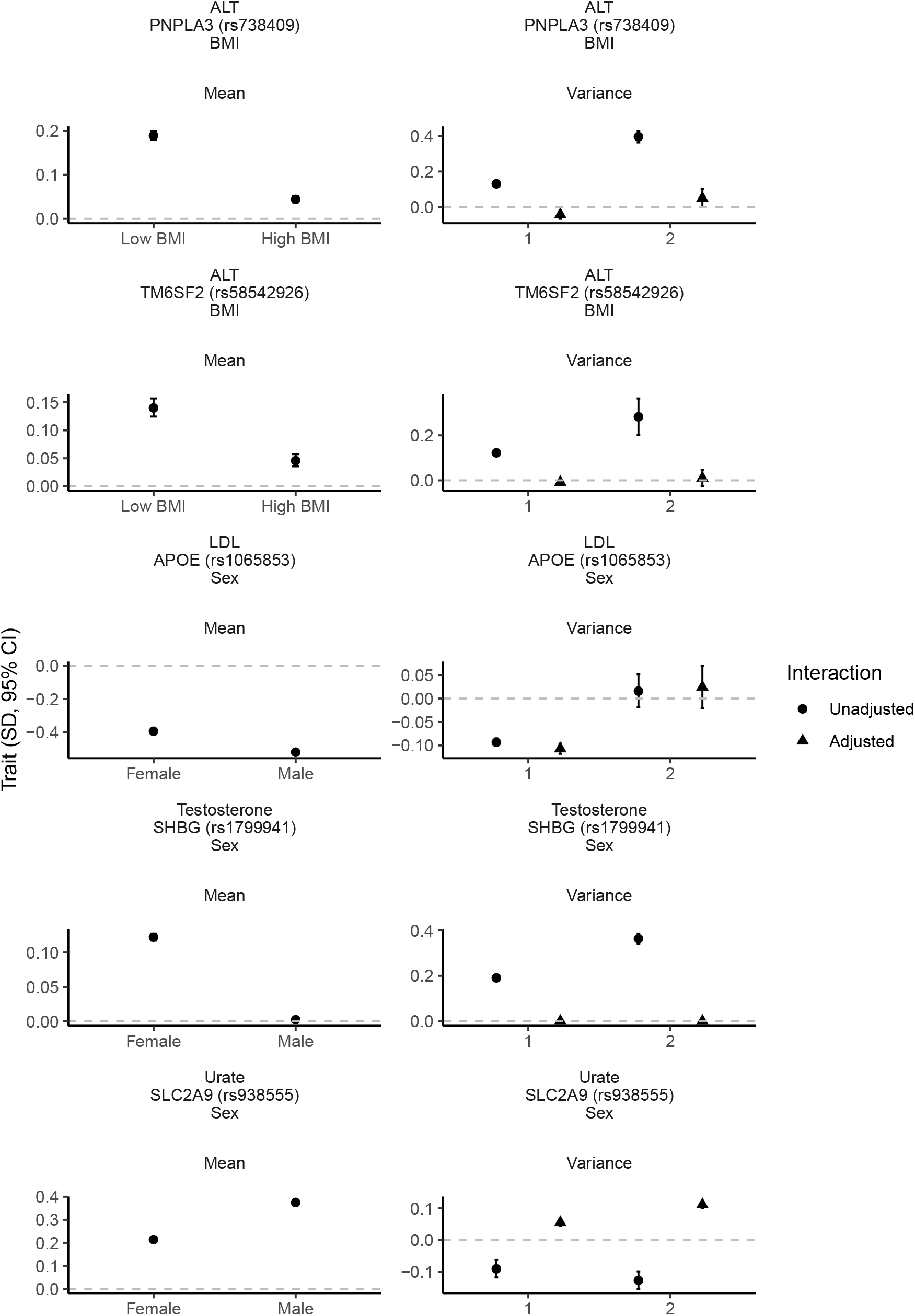
Effect of top gene-environment interaction loci on trait mean and variance. Per-allele effect of SNP stratified by modifier on outcome mean estimated with heteroscedastic-consistent standard errors^29^ and unstratified effect of SNP on variance estimated using LAD-BF (genotype 0 vs 1 and 0 vs 2) with or without adjustment for the interaction term. All estimates were adjusted for age, sex (except for rs1065853, rs1799941 and rs938555 on variance as the modifier was sex) and top ten genetic principal components. SD, standard deviation. CI, confidence interval. ALT, alanine aminotransferase. LDL, low-density lipoprotein. BMI, body mass index. Low BMI, <= 26.7 kg/m^2^. High BMI, > 26.7 kg/m^2^.

### Gene-gene interaction effects (GxG)

We detected eight GxG effects on the additive scale (P < 5 × 10^−8^; **Figure S12**), six of which were also associated on the multiplicative scale (P < 5 × 10^−8^; **Figure S13**). There was no strong attenuation following adjustment for fine-mapped main effects (**Figure S14**) suggesting phantom epistasis^41,47^ was not a major source of bias. *ZNF827* rs4835265 x *NEDD4L* rs4503880 was inversely associated with -0.04 SD (95% CI -0.05, -0.03) gamma glutamyltransferase (GGT), *ABO* rs635634 x *FUT2* rs281379, *ABO* rs635634 x *TREH* rs12225548, and *TREH* rs12225548 x *FUT2* rs281379 were associated with 0.08 SD (95% CI 0.07, 0.09), 0.04 SD (95% CI 0.03, 0.05) and 0.02 SD (95% CI 0.02, 0.03) increase in alkaline phosphatase (ALP) respectively, *HSD17B13* rs71633359 *x PNPLA3* rs738409 and *HSD17B13* rs71633359 x *PNPLA3* rs3747207 were associated with -0.04 SD (95% CI -0.05, -0.03) and - 0.04 SD (95% CI -0.05, -0.03) decrease in ALT and aspartate aminotransferase (AST) respectively (**Figure 3**). Adjusting the variance effects for the interaction term had no strong impact on the variance estimate (**Figure 3**).

**Figure 3.**
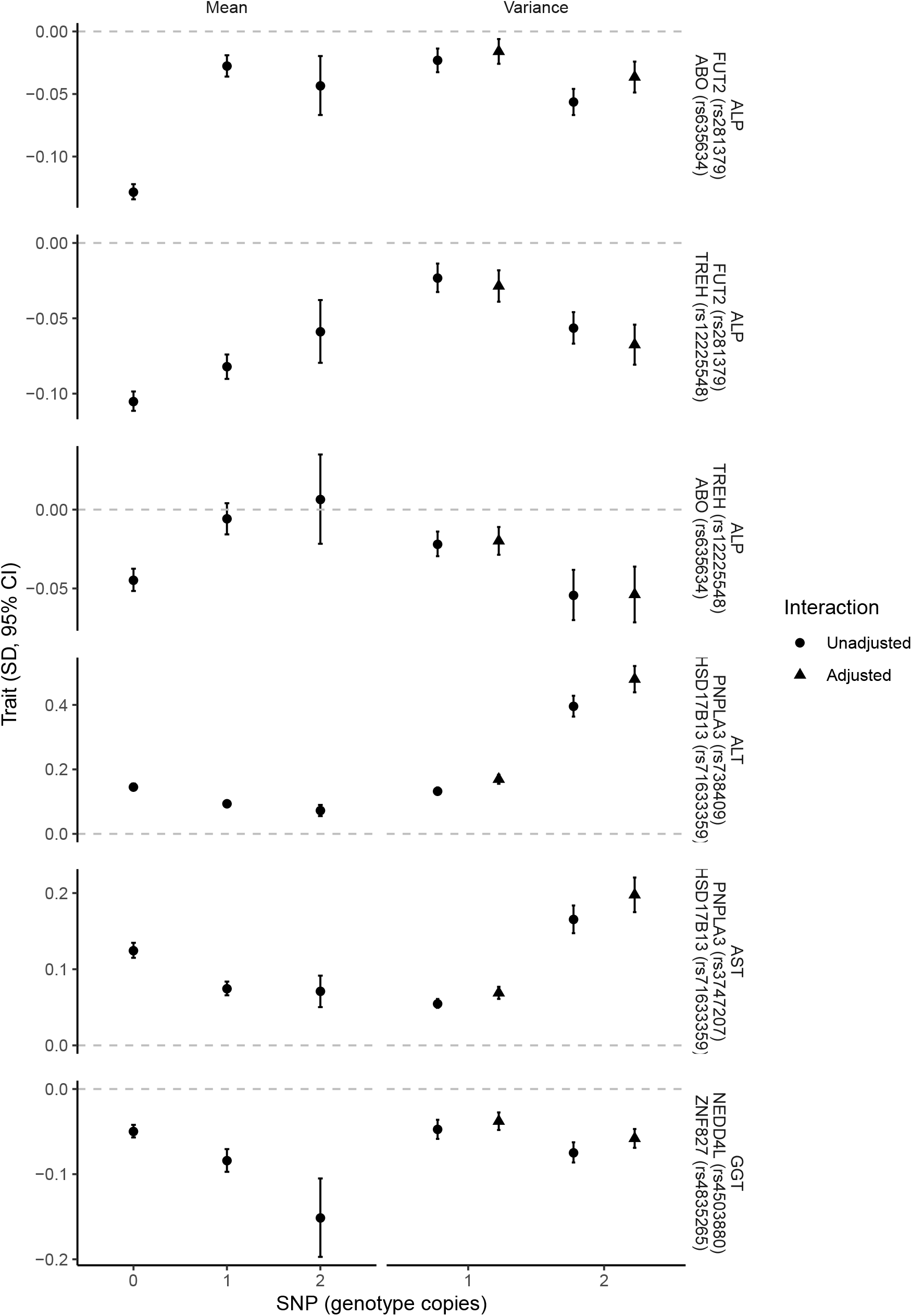
Effect of top gene-gene interaction loci on trait mean and variance. Per allele effect of SNP stratified by modifier on outcome mean estimated with heteroscedastic-consistent standard errors^29^ and unstratified effect of SNP on variance estimated using LAD-BF (genotype 0 vs 1 and 0 vs 2) with or without adjustment for the interaction term. All estimates were adjusted for age, sex, and top ten genetic principal components. SD, standard deviation. CI, confidence interval. ALP, alkaline phosphatase. ALT, alanine aminotransferase. AST, aspartate aminotransferase. GGT, gamma glutamyltransferase.

## Discussion

Here we demonstrate the value of variance GWAS in identifying 468 independent vQTLs with evidence of interaction on 24 serum biochemistry phenotypes in UK Biobank and subsequently identify 82 GxE and six GxG scale independent effects. To facilitate this large-scale analysis on∼337K UK Biobank participants we developed an efficient C++ implementation of a LAD regression-based Brown-Forsythe test^22^ (implemented in varGWAS) with functionality to reliably estimate variance effects and compared the test with the original non-parametric version (implemented in OSCA^13,26^) through a series of simulations.

Although the power to detect genetic interaction effects using variance prioritisation was low, when applied to large sample sizes such as UK Biobank strong evidence for association can be identified as demonstrated in this study and by Wang *et al*^13^. We found LAD-BF had several advantages over the original non-parametric test when applied to GWAS. First, LAD-BF directly supports adjustment for covariates (although this could be achieved using the original test if applied to pre-adjusted phenotypes^13^). Second, LAD-BF can test effects of continuous genotypes which enables application to the expected genotype value (“dose”) from imputed SNP array data. Third, our model provides a variance effect estimate which is valid when there is a SNP interaction effect, unlike the implementation of the original Brown-Forsythe test in OSCA which provides an incorrect variance effect estimate derived from the test P-value^27^. We also demonstrate through simulation that adjusting the variance effect for the interaction term causes attenuation which is useful to determine if other interactions exist and could potentially be applied using stepwise regression until all interaction effects are discovered, subject to sufficient power. However, there are some disadvantages. The runtime was 75% longer than the original test implemented in OSCA, although this is still fast enough to allow large-scale analyses. Second, the effect estimate (but not test statistic) is based on normality assumptions which may be violated in practice.

The largest GxE effects replicate existing findings: *PNPLA3* rs738409 x BMI on ALT levels^48,49^, *SLC2A9* rs938555 x sex on urate^50^, *APOE* rs1065853 x sex on LDL^51^, *SHBG* rs1799941 x sex on testosterone^52^, and *TM6SF2* rs58542926 x BMI on ALT^48^. Adjusting the variance effect for the interaction led to attenuation of *PNPLA3* rs738409 and *TM6SF2* rs58542926 on ALT and *SHBG* rs1799941 on testosterone, however strong evidence of variance effects remained for ALT at *PNPLA3* rs738409 and *TM6SF2* rs58542926 suggesting other interaction effects may exist at these loci. The variance effect of *SHBG* rs1799941 on testosterone was weak after adjusting for rs1799941 x sex suggesting no strong evidence of further interaction effects on testosterone at this locus, but the test may be underpowered to detect additional effects.

We replicated previous GxG effects of *ABO* rs635634 x *FUT2* rs281379 on ALP^53,54^ and *HSD17B13* rs71633359 x *PNPLA3* rs738409/rs3747207 on ALT and AST^55,56^ and find no strong evidence of ‘phantom epistasis’ ^41,47^ as a potential explanation. Additionally, we identified novel effects of *TREH* rs12225548 x *FUT2* rs281379 and *ABO* rs635634 x *TREH* rs12225548 on ALP and *ZNF827* rs4835265 x *NEDD4L* rs4503880 on GGT. ABO blood group antigens and secretion status are thought to influence ALP clearance^57,58^. *TREH* rs12225548 has a strong main effect on ALP^39,59,60^ and interactions of these loci may be explained by interplay of ALP production and clearance mechanisms. *ZNF827* and *NEDD4L* loci have previously been reported to influence GGT levels in independent populations but the mechanism is unclear^61,62^.

None of the GxG loci variance effects strongly attenuated after adjusting for the interaction term. This could be a consequence of low power since the interaction effect likely explains a very small amount of the trait variance but could also indicate the presence of other interaction effects involving the same SNP not included in the variance model. Indeed, we found strong GxE evidence at some of these loci: *ABO* rs635634 x sex on ALP, *HSD17B13* rs71633359 x BMI and *PNPLA3* rs738409/rs3747207 x BMI on ALT and AST.

Evidence of gene-interaction effects could suggest the protein product also has an interaction effect. In which case interventions developed to target the protein will show differential effects on the indication and could have low or no efficacy in some subgroups^63^. Such evidence could be important to support developments in stratified medicine. Therefore, vQTL evidence may have a role in preclinical drug development to deprioritise targets given the possibility of target-outcome heterogeneous effects. Further research is needed to appraise the utility of vQTLs in the drug development pipeline as has been done for protein QTLs^64^.

However, there are other explanations for vQTLs that are not in terms of biology. First, loci that are weakly correlated with a SNP having a strong main effect can introduce a phantom vQTL^65,66^. In this situation variance is introduced through variability in LD between the artefactual vQTL and QTL. Second, vQTLs could signify fluctuation of a trait measurement within an individual over time^10^ and may originate from normal biological processes such as circadian rhythm. Third, we assume homogeneity of variance within each genotype group which could be violated by the mean-variance relationship and observed low concordance of vQTL effects on the log and natural scales are evidence for this. Additionally, our interactions could be explained by non-linear relationships between the exposure and outcome or scale artefacts^67^. We sought to reduce the latter by replicating effects on additive and multiplicative scales.

Through this work we performed hypothesis-free analyses of genetic interaction effects on 30 blood biomarkers in UK Biobank using variance prioritisation and found evidence for 88 effects. Many of our top findings replicate previously reported associations, but we also report first evidence of *TREH* rs12225548 x *FUT2* rs281379 and *TREH* rs12225548 x *ABO* rs635634 on ALP and *ZNF827* rs4835265 x *NEDD4L* rs4503880 on GGT. Additionally, we show variance attenuation of *PNPLA3* rs738409 and *TM6SF2* rs58542926 on ALT and *SHBG* rs1799941 on testosterone after adjusting for the interaction indicating these effects were contributing to the variance association, but the ALT effects were still strong suggesting additional interactions may exist at these loci. These data could be used to discover possible subgroup effects for a given biomarker during preclinical drug development. To facilitate our analysis, we developed C++ variance GWAS software that implements a LAD-regression based Brown-Forsythe test, provide a convenient R-package based on this software and introduce methodology to estimate the variance effects which can be applied to other studies.

## Data Availability

Software to perform variance GWAS using the LAD Brown-Forsythe model is available from https://github.com/MRCIEU/varGWAS and R-package for ad hoc analyses is available from https://github.com/MRCIEU/varGWASR. Code for performing simulation studies is available from https://github.com/MRCIEU/varGWAS/sim. Code for running the UK Biobank analysis is available from https://github.com/MRCIEU/varGWAS-ukbb-biomarkers. Full variance GWAS summary statistics are available from the OpenGWAS project. All code repositories are available under the GPL v3 license.

https://github.com/MRCIEU/varGWASR

https://github.com/MRCIEU/varGWAS-ukbb-biomarkers

https://gwas.mrcieu.ac.uk/

## Supplemental information description

Supplemental Material and Methods

Figure S1. UK Biobank participant inclusion criteria

Figure S2. Power to detect SNP-interaction effects using variance testing under simulation

Figure S3. Type I error of Brown-Forsythe tests

Figure S4. Effect of adjustment for the interaction effect on variance test P-value distribution

Figure S5. Runtime performance of varGWAS and OSCA

Figure S6. Manhattan plots of biomarker variance GWAS using regression-based Brown-Forsythe test

Figure S7. Q-Q plots of biomarker variance GWAS using regression-based Brown-Forsythe test

Figure S8. Biomarker distribution

Figure S9. Top gene-by-environment interaction effects (P < 5 × 10^−8^) on biomarker concentration using additive scale

Figure S10. Top gene-by-environment interaction effects (P < 5 × 10^−8^) on biomarker concentration using multiplicative scale

Figure S11. Top gene-by-environment interaction effects (P < 5 × 10^−8^) on biomarker concentration using additive scale adjusted for fine-mapped main effect

Figure S12. Top gene-by-gene interaction effects (P < 5 × 10^−8^) on biomarker concentration using additive scale

Figure S13. Top gene-by-gene interaction effects (P < 5 × 10^−8^) on biomarker concentration using multiplicative scale

Figure S14. Top gene-by-gene interaction effects (P < 5 × 10^−8^) on biomarker concentration using additive scale adjusted for fine-mapped main effects

Table S1. GWAS summary statistics for top vQTLs identified through this study

Table S2. Top GxG/GxE effect summary statistics

Table S3. Fine-mapped loci covariates

## Acknowledgements

This study was funded by the NIHR Biomedical Research Centre at University Hospitals Bristol and Weston NHS Foundation Trust and the University of Bristol. The views expressed are those of the author(s) and not necessarily those of the NIHR or the Department of Health and Social Care. This work was also funded by the UK Medical Research Council as part of the MRC Integrative Epidemiology Unit (MC_UU_00011/1, MC_UU_00011/3 and MC_UU_00011/4). L.A.C.M is funded by a University of Bristol Vice-Chancellor’s fellowship.

## Code and data availability

Software to perform variance GWAS using the LAD Brown-Forsythe model is available from https://github.com/MRCIEU/varGWAS and R-package for *ad hoc* analyses is available from https://github.com/MRCIEU/varGWASR. Code for performing simulation studies is available from https://github.com/MRCIEU/varGWAS/sim. Code for running the UK Biobank analysis is available from https://github.com/MRCIEU/varGWAS-ukbb-biomarkers. Full variance GWAS summary statistics are available from the OpenGWAS project^39,68^. All code repositories are available under the GPL v3 license.

