## Supplemental Text for "Hypothesis-free detection of gene-interaction effects on biomarker concentration in UK Biobank using variance prioritisation"

### Supplemental Material and Methods

#### Simulation studies

##### Simulation to estimate the accuracy of variance effect estimate and confidence interval coverage (Figure 1)

*Aim*^1^: To qualitatively evaluate the accuracy of variance effect estimates of original Brown-Forsythe test with effect size derived from P-value^2^ and regression-based Brown-Forsythe tests under linear variance effect or interaction of genotype-by-continuous modifier.

*Data-generating mechanisms*: Data were simulated for N=10,000 independent observations within each simulated dataset. For each observation, we simulated a genotype in Hardy-Weinberg equilibrium (HWE) with a minor allele frequency (MAF) of 0.4 and standard Normal outcome. The genotype was set to have linear effect on outcome variance or an interaction effect. The effects were set as in Brookes *et al*^3^ with the main effect fixed across all simulations and set to have 95% power. The magnitude of the variance/interaction effect size was varied and set relative to the main effect ranging from 0x to 12x in 1x increments. The outcome was scaled to have zero mean and unit variance.

*Estimand*: The effect of variance difference between genotype 0 and 1 and 0 and 2. The proportion of replicates where the effect estimate 95% confidence included the true variance difference.

*Methods*: The effect of simulated genotype on outcome variance was measured using the original and regression-based Brown-Forsythe tests (see main methods). Each configuration of parameters was evaluated using N=1000 replications.

*Performance measures*: Accuracy of effect size estimates. Coverage of 95% confidence intervals.

*Open-source code*: https://github.com/MRCIEU/varGWAS/blob/master/sim/sim12.R

##### Simulation to estimate the power of variance tests to detect interaction effects (Figure S2)

*Aim*^1^: To estimate and compare statistical power of original and regression-based Brown-Forsythe tests to detect variance effects produced by simulated interaction of genotype-by-continuous modifier on continuous outcomes.

*Data-generating mechanisms*: Data were simulated for N=200, N=2000, N=200,000 and N=2,000,000 independent observations within each simulated dataset. For each observation, we simulated a genotype in HWE with a MAF of 0.4, a standard Normal modifier and residual drawn from either standard Normal, t (df=4) or log standard Normal distributions. The effects were set as in Brookes *et al*^3^ with the main effect fixed across all simulations and set to have 80% power when the sample size was N=200 (assuming normally distributed residuals). The interaction effect size was varied and set relative to the main effect ranging from 0x to 6x in 0.5x increments.

*Estimand*: The P value for the null hypothesis of variance homogeneity.

*Methods*: The effect of simulated genotype on outcome variance was measured using original and regression-based Brown-Forsythe tests (see main methods). Each configuration of parameters was evaluated using N=200 replications.

*Performance measures*: Power was defined as the percentage of tests with P < 0.05.

*Open-source code*: https://github.com/MRCIEU/varGWAS/blob/master/sim/sim1.R

##### Simulation to estimate type I error of variance tests under the null hypothesis (Figure S3)

*Aim*^1^: To qualitatively evaluate and compare type I error of original and regression-based Brown-Forsythe tests under the null effect of a simulated genotype on continuous outcomes.

*Data-generating mechanisms*: Data were simulated for N=100,000 independent observations within each simulated dataset. For each observation, we simulated genotypes in HWE with MAF of 0.05 and a residual drawn from either standard Normal, t (df=4), log standard Normal or mixed Normal $0.9 N\left( 0,1 \right), 0.1 N(5,1)$ distributions. The effect of the genotype on outcome was set to null.

*Estimand*: The P value for the null hypothesis of variance homogeneity.

*Methods*: The effect of the genotype on outcome variance was tested using the original and regression-based Brown-Forsythe tests (see main methods) with N=1000 replications.

*Performance measures*: Q-Q plot of the test P values and expected null distribution.

*Open-source code*: https://github.com/MRCIEU/varGWAS/blob/master/sim/sim2b.R

##### Simulation to compare P-value distributions of regression-based Brown-Forsythe test with and without adjustment for the interaction effect (Figure S4)

*Aim*^1^: To qualitatively evaluate and compare the P-value distribution of regression-based Brown-Forsythe test under interaction of simulated genotype on continuous outcome with/without adjustment for interaction effect.

*Data-generating mechanisms*: Data were simulated for N=1000 independent observations within each simulated dataset. For each observation, we simulated a genotype in HWE with MAF of 0.4. The SNP was simulated to have a main effect and interaction effect explaining 6.5% and 20% of the variance of the outcome, respectively. The outcome was drawn from standard Normal distribution.

*Estimand*: The P value for the null hypothesis of variance homogeneity.

*Methods*: The effect of the genotype on outcome variance was tested using regression-based Brown-Forsythe test (see main methods) with/without adjustment for the modifier and interaction term in the first-stage regression model with N=1000 replications.

*Performance measures*: Q-Q plot of the test P values and expected null distribution.

*Open-source code*: https://github.com/MRCIEU/varGWAS/blob/master/sim/sim13.R

##### Simulation to estimate runtime performance of variance tests (Figure S5)

*Aim*^1^: To estimate and compare runtime performance of original and regression-based Brown-Forsythe tests with no effect of a simulated genotype on continuous outcome with increasing CPU threads

*Data-generating mechanisms*: Data were simulated for N=100,000 independent observations within each simulated dataset. For each observation, we simulated 1000 independent genotypes in HWE with a MAF of 0.4 and a residual drawn from standard Normal distribution.

*Estimand*: The execution runtime in seconds.

*Methods*: The effect of the genotype on outcome variance was tested using non-parametric and regression-based Brown-Forsythe tests (see main methods) with N=200 replications and with increasing CPU threads: 1, 2, 4 and 8 using an Intel(R) Xeon(R) CPU E5-2680 v4 @ 2.40GHz.

*Performance measures*: Plot of runtime between methods.

*Open-source code*: https://github.com/MRCIEU/varGWAS/blob/master/sim/sim4.R

#### Fine mapping of main effect SNPs

Fine-mapping was performed between natural linkage disequilibrium break points identified in European populations^4^ containing the interacting variant using SuSiE^5^ assuming at most 10 casual variants. The data were processed using gwasglue R-package^6^. Summary statistics for the SNP-biomarker main effects were obtained from Neale *et al*^6,7^. European 1000 genome phase 3 linkage disequilibrium matrices were obtained from OpenGWAS containing bi-allelic SNPs with MAF > 0.01^6^.

**Figure S1. UK Biobank participant inclusion criteria**

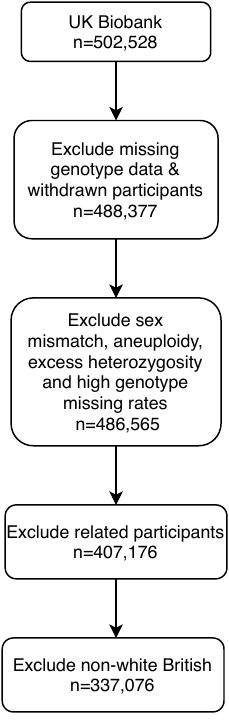

**Figure S2. Power to detect SNP-interaction effects using variance testing under simulation**

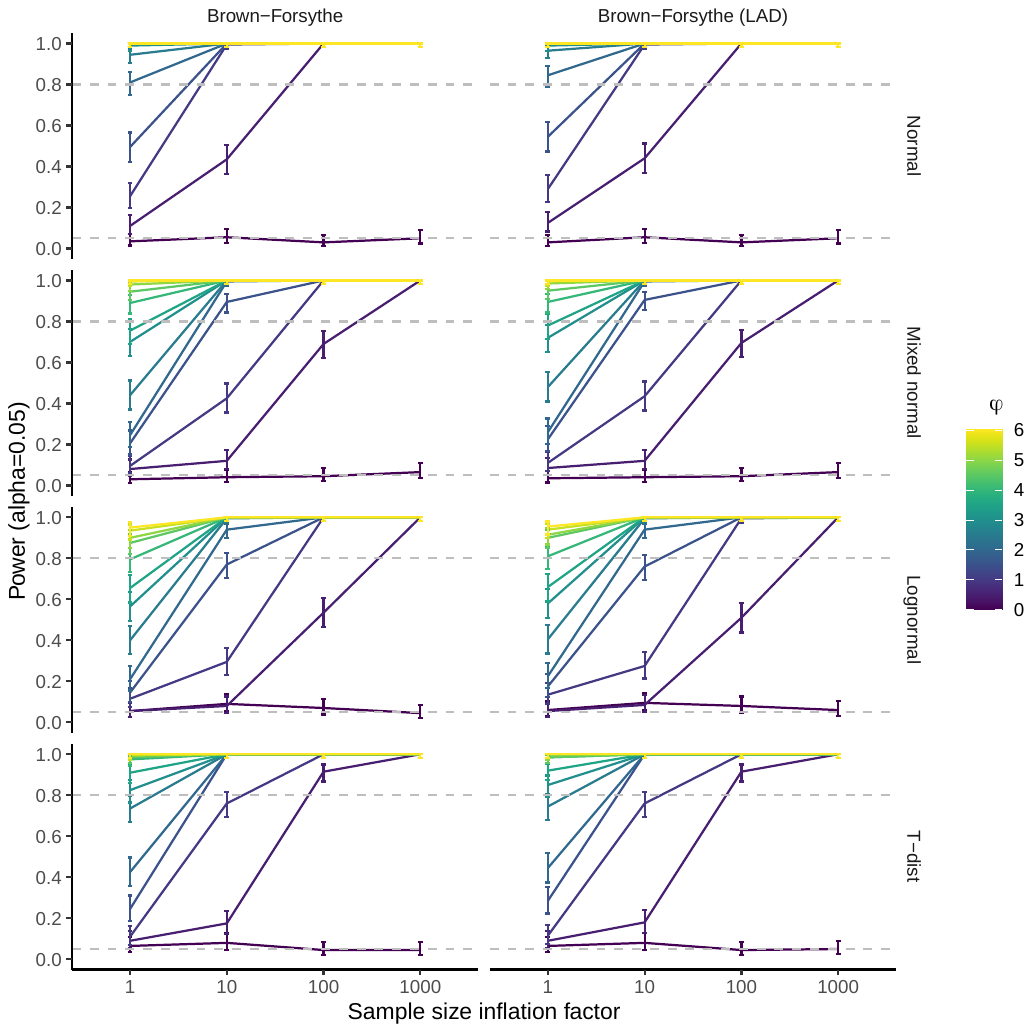

Φ, Interaction effect size relative to main effect. Inflation factor, sample size relative to the size required to detect the main effect with 80% power. Normal, distribution with mean of 0 and variance of 1. Mixed normal, distribution with 90% Normal with mean of 0 and variance of 1 and 10% Normal with mean of 5 and variance of 1. Lognormal, distribution with mean of 0 and variance of 1. T-dist, distribution with 4 degrees of freedom. SNP, single-nucleotide polymorphism simulated with minor allele frequency of 0.4 in Hardy-Weinberg equilibrium. All simulations had a fixed main effect detectable with 80% power when the sample size inflation factor was equal to 1. Simulation was produced with 200 repetitions. Sample size inflation factor of 1 was set to 200 observations. Error bars represent the 95% confidence interval. LAD, least absolute deviation.

**Figure S3. Type I error of Brown-Forsythe tests**

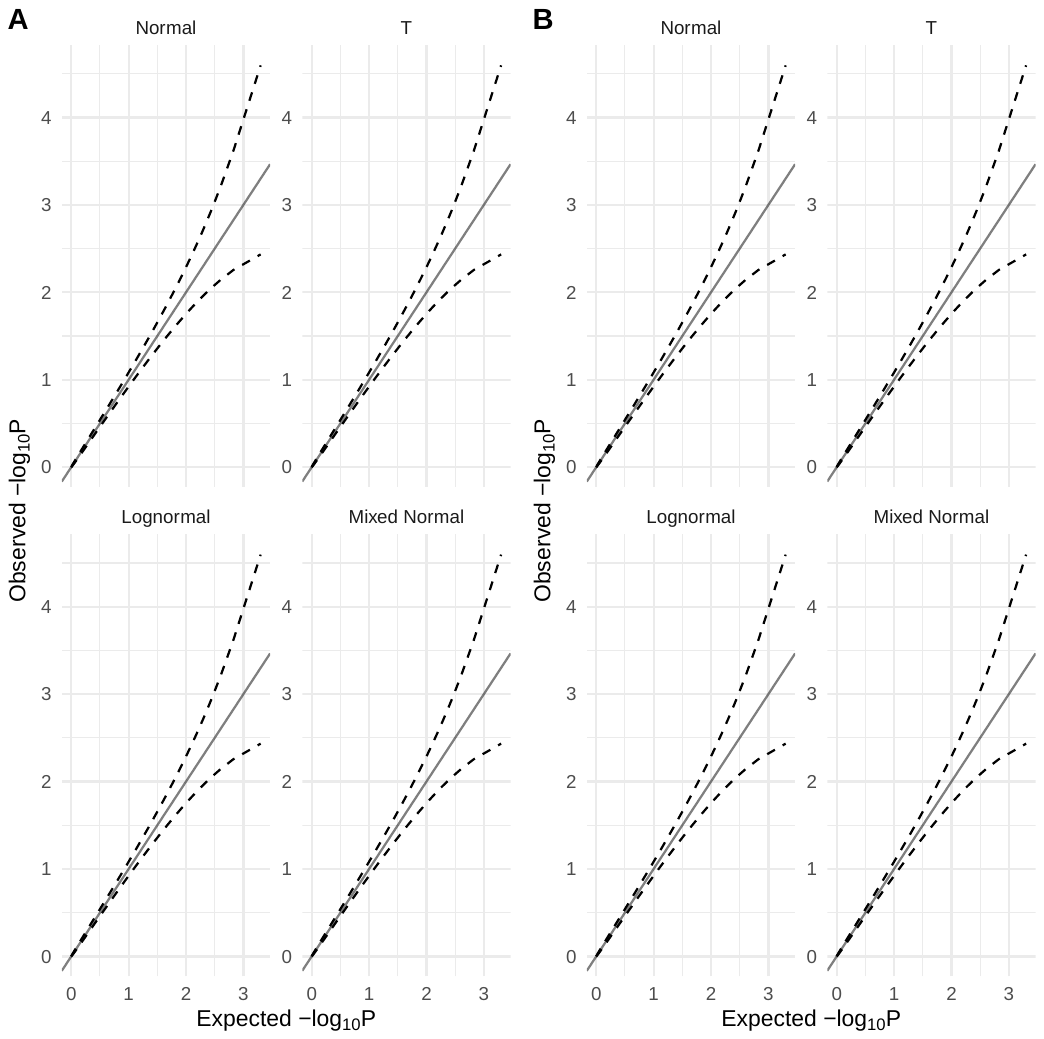

Normal, standard Normal distribution. Lognormal, standard log Normal distribution. T-dist, distribution with 4 degrees of freedom. Mixed normal, distribution produced with $0.9 N\left( 0,1 \right), 0.1 N(5,1)$. A, LAD regression Brown-Forsythe. B, Original Brown-Forsythe test. Simulations were produced with 1000 repetitions and 100,000 observations. Dashes represent the 95% confidence interval.

**Figure S4. Effect of adjustment for the interaction effect on variance test P-value distribution**

| A  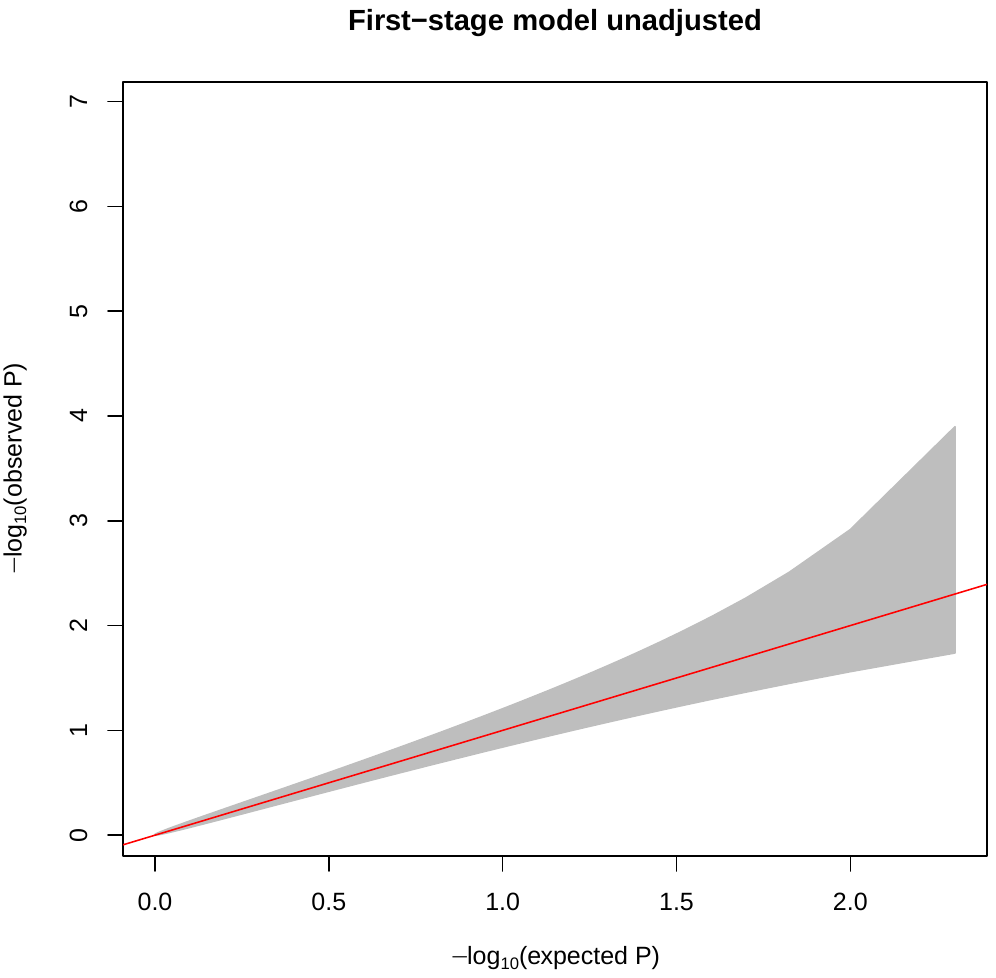 | B  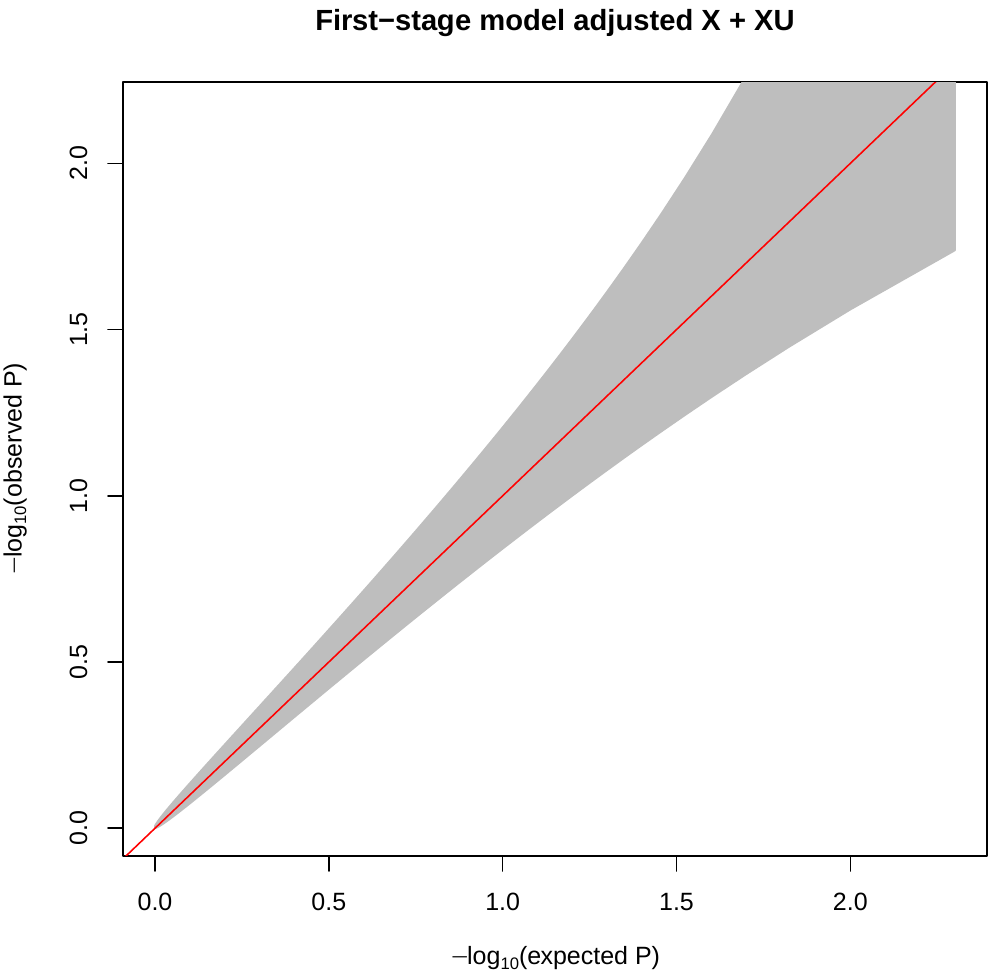 |
| --- | --- |

SNP simulated to have a main and interaction effect explaining 6.5% and 20% of the variance of a standard Normal outcome, respectively. The SNP was tested for a variance effect using regression-based Brown-Forsythe test. A, No adjustment. B, Adjustment for interaction in the first-stage regression model.

**Figure S5. Runtime performance of varGWAS and OSCA**

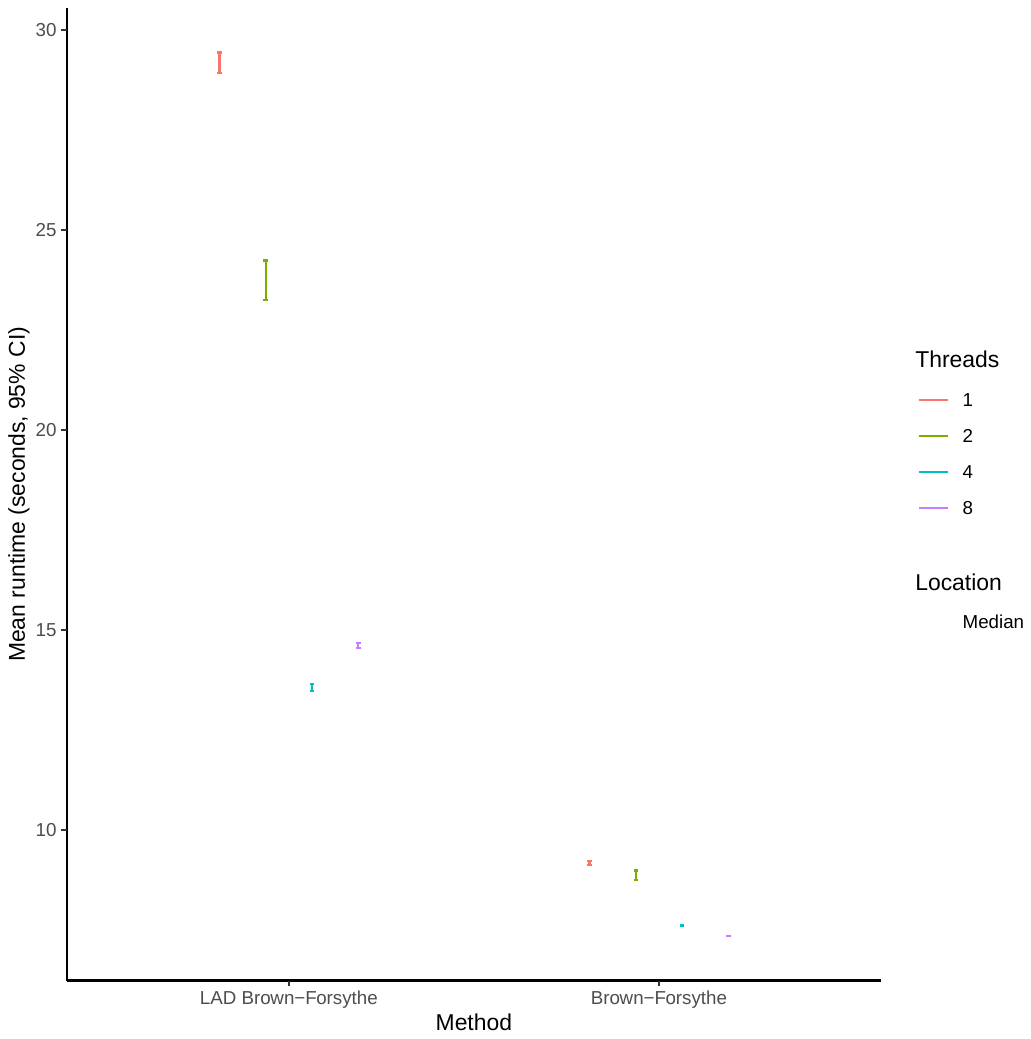

Average runtime for effect of 1000 SNPs tested on outcome variance using LAD-BF implemented in varGWAS and original Brown-Forsythe implemented in OSCA with increasing CPU threads. CI, confidence interval.

**Figure S6. Manhattan plots of biomarker variance GWAS using regression-based Brown-Forsythe test**

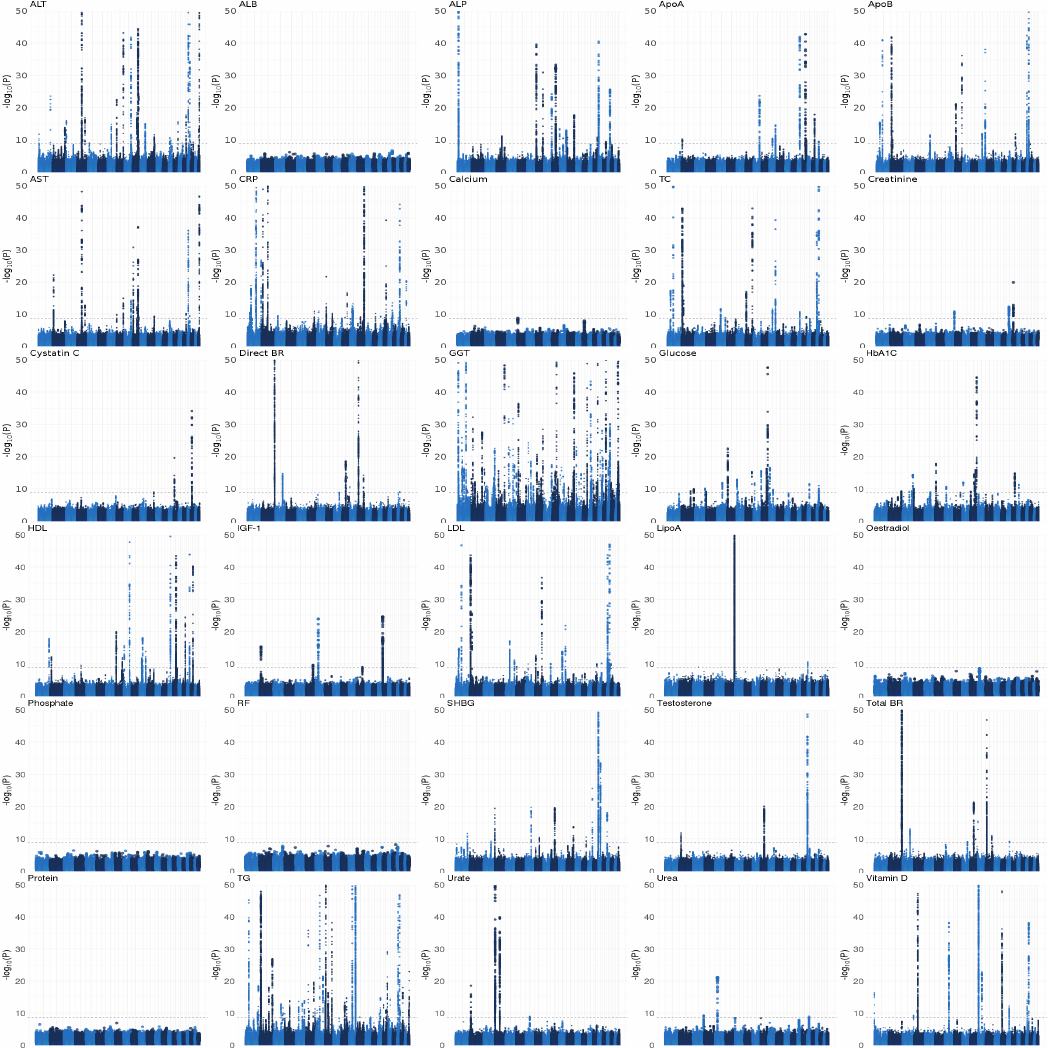

Manhattan plots for GWAS of SNP effects on biomarker variability adjusted for age, sex, and top ten genetic principal components. ALB, albumin. ALP, alkaline phosphatase. ALT, alanine aminotransferase. AST, aspartate aminotransferase. ApoA, Apolipoprotein A. ApoB, apolipoprotein B. CRP, c-reactive protein. Direct BR, direct bilirubin. GGT, Gamma glutamyltransferase. HDL, high-density lipoprotein. HbA1C, glycated haemoglobin. IGF-1, insulin growth factor. LDL, low-density lipoprotein. LipoA, lipoprotein A. RF, rheumatic factor. SHBG, sex-hormone binding globulin. TC, total cholesterol. TG, triglycerides. Total BR, total bilirubin. Biomarker outliers with Z-score > 5SD from the mean were removed to control type I error.

**Figure S7. Q-Q plots of biomarker variance GWAS using regression-based Brown-Forsythe test**

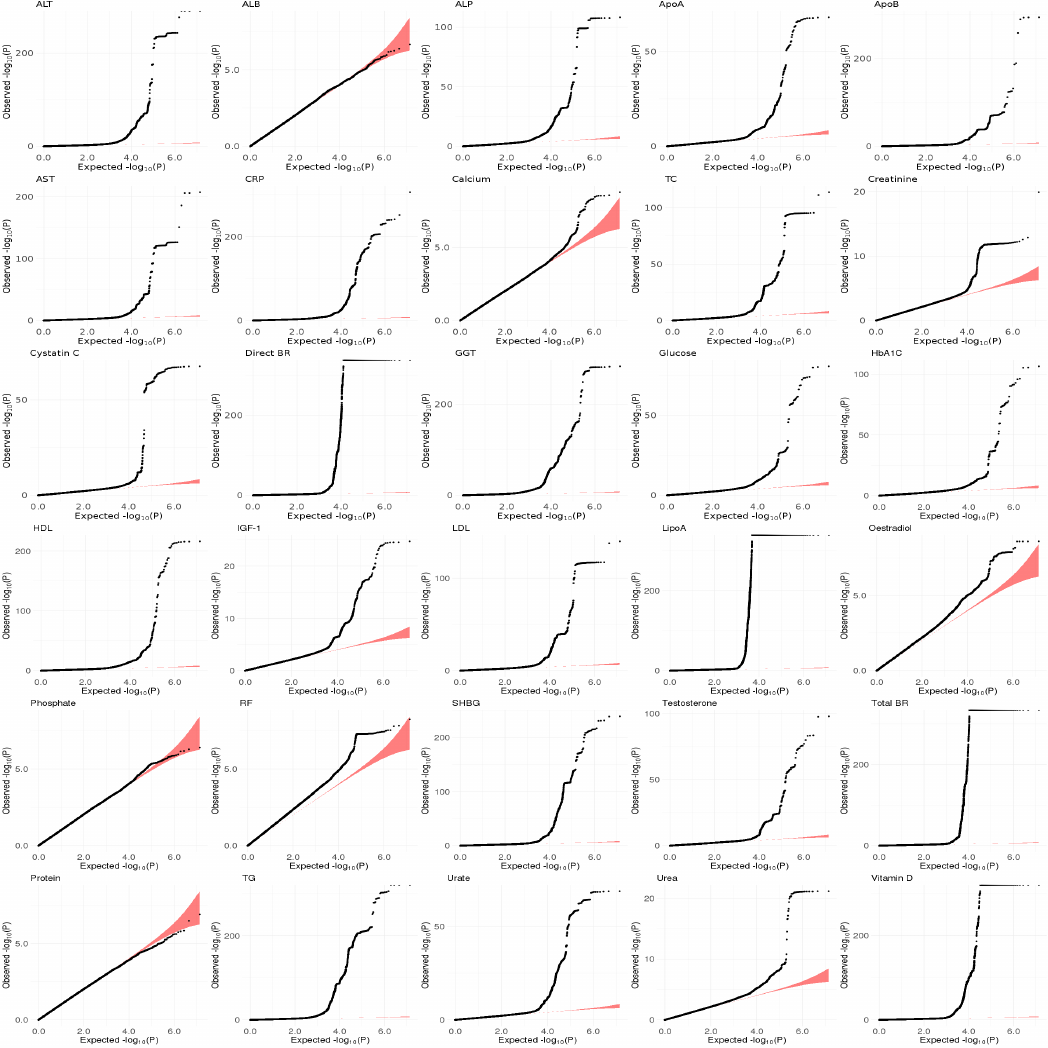

Q-Q plots for GWAS of SNP effects on biomarker variance adjusted for age, sex, and top ten genetic principal components. ALB, albumin. ALP, alkaline phosphatase. ALT, alanine aminotransferase. AST, aspartate aminotransferase. ApoA, Apolipoprotein A. ApoB, apolipoprotein B. CRP, c-reactive protein. Direct BR, direct bilirubin. GGT, Gamma glutamyltransferase. HDL, high-density lipoprotein. HbA1C, glycated haemoglobin. IGF-1, insulin growth factor. LDL, low-density lipoprotein. LipoA, lipoprotein A. RF, rheumatic factor. SHBG, sex-hormone binding globulin. TC, total cholesterol. TG, triglycerides. Total BR, total bilirubin. Biomarker outliers with Z-score > 5SD from the mean were removed to control type I error.

**Figure S8. Biomarker distribution**

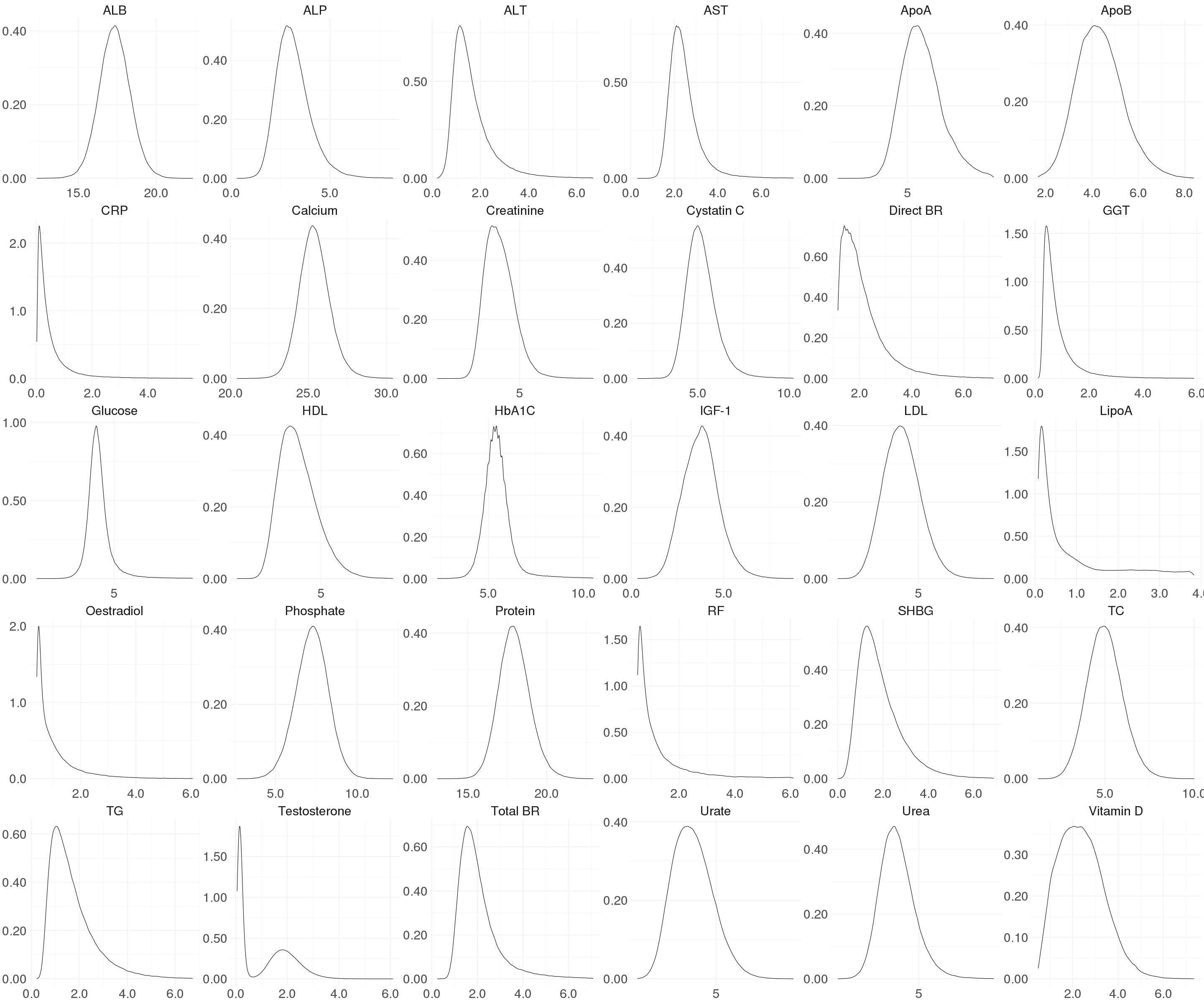

ALB, albumin. ALP, alkaline phosphatase. ALT, alanine aminotransferase. AST, aspartate aminotransferase. ApoA, Apolipoprotein A. ApoB, apolipoprotein B. CRP, c-reactive protein. Direct BR, direct bilirubin. GGT, Gamma glutamyltransferase. HDL, high-density lipoprotein. HbA1C, glycated haemoglobin. IGF-1, insulin growth factor. LDL, low-density lipoprotein. LipoA, lipoprotein A. RF, rheumatic factor. SHBG, sex-hormone binding globulin. TC, total cholesterol. TG, triglycerides. Total BR, total bilirubin. SD units. Biomarker outliers with Z-score > 5SD from the mean were removed.

**Figure S9. Top gene-by-environment interaction effects (P < 5 x 10^-8^) on biomarker concentration using additive scale**

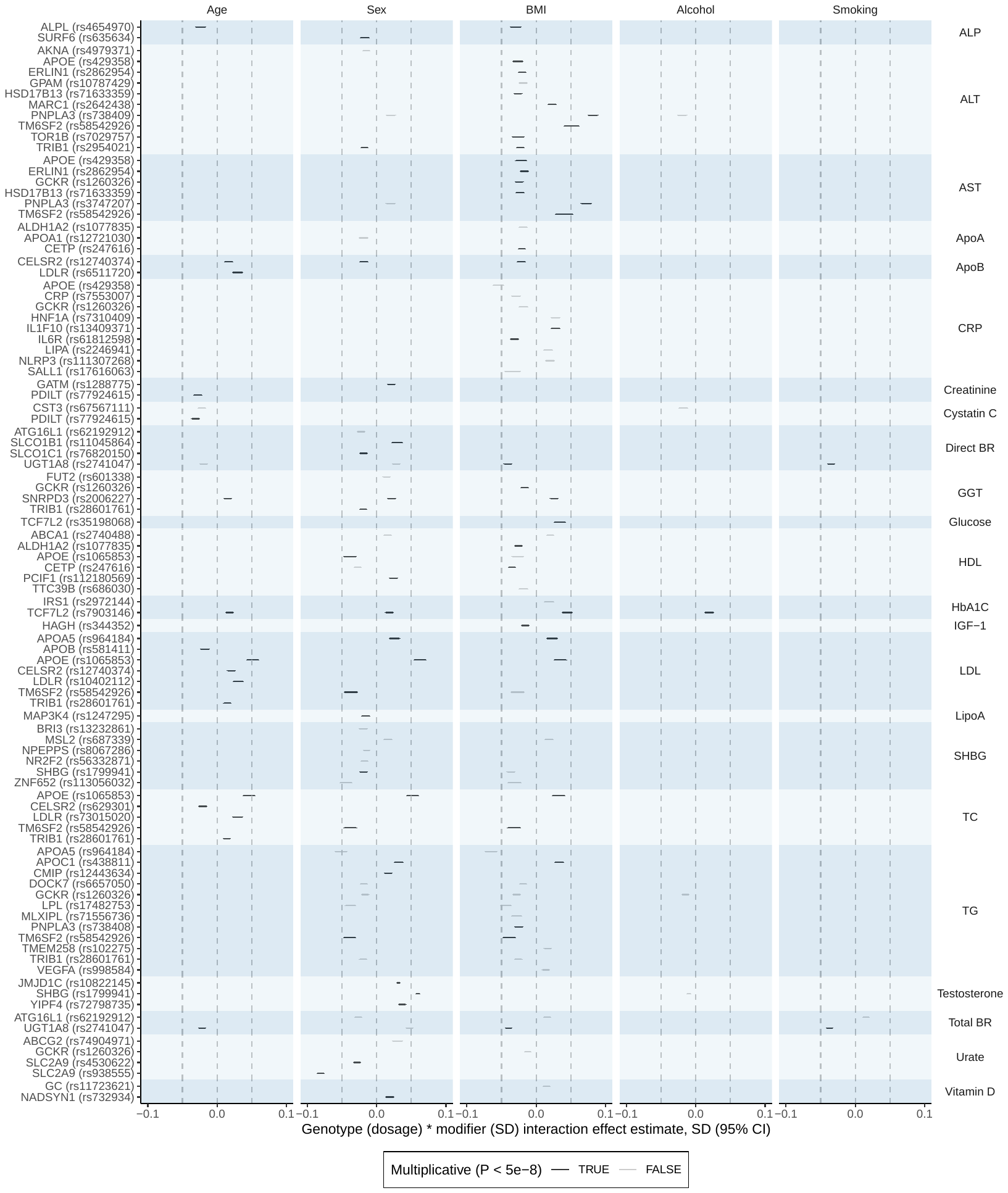

GxE effects using additive scale and heteroscedasticity consistent standard errors^8^ (P < 5 x 10^-8^). ALP, alkaline phosphatase. ALT, alanine aminotransferase. AST, aspartate aminotransferase. ApoA, Apolipoprotein A. ApoB, apolipoprotein B. CRP, C-reactive protein. Direct BR, direct bilirubin. GGT, Gamma glutamyltransferase. HDL, high-density lipoprotein. HbA1c, glycated haemoglobin. IGF-1, insulin-like growth factor 1. LDL, low-density lipoprotein. LipoA, lipoprotein A. SHBG, sex-hormone binding globulin. TC, total cholesterol. TG, triglycerides. Total BR, total bilirubin. BMI, body mass index. Smoking, smoking status. Alcohol, intake. PA, physical activity. All measures reported on SD scale. All estimates were adjusted for the main effect, age, sex, and top ten genetic principal components. Gene name is the nearest protein coding gene HGNC name by chromosomal position. SD, standard deviation. CI, confidence interval.

**Figure S10. Top gene-by-environment interaction effects (P < 5 x 10^-8^) on biomarker concentration using multiplicative scale**

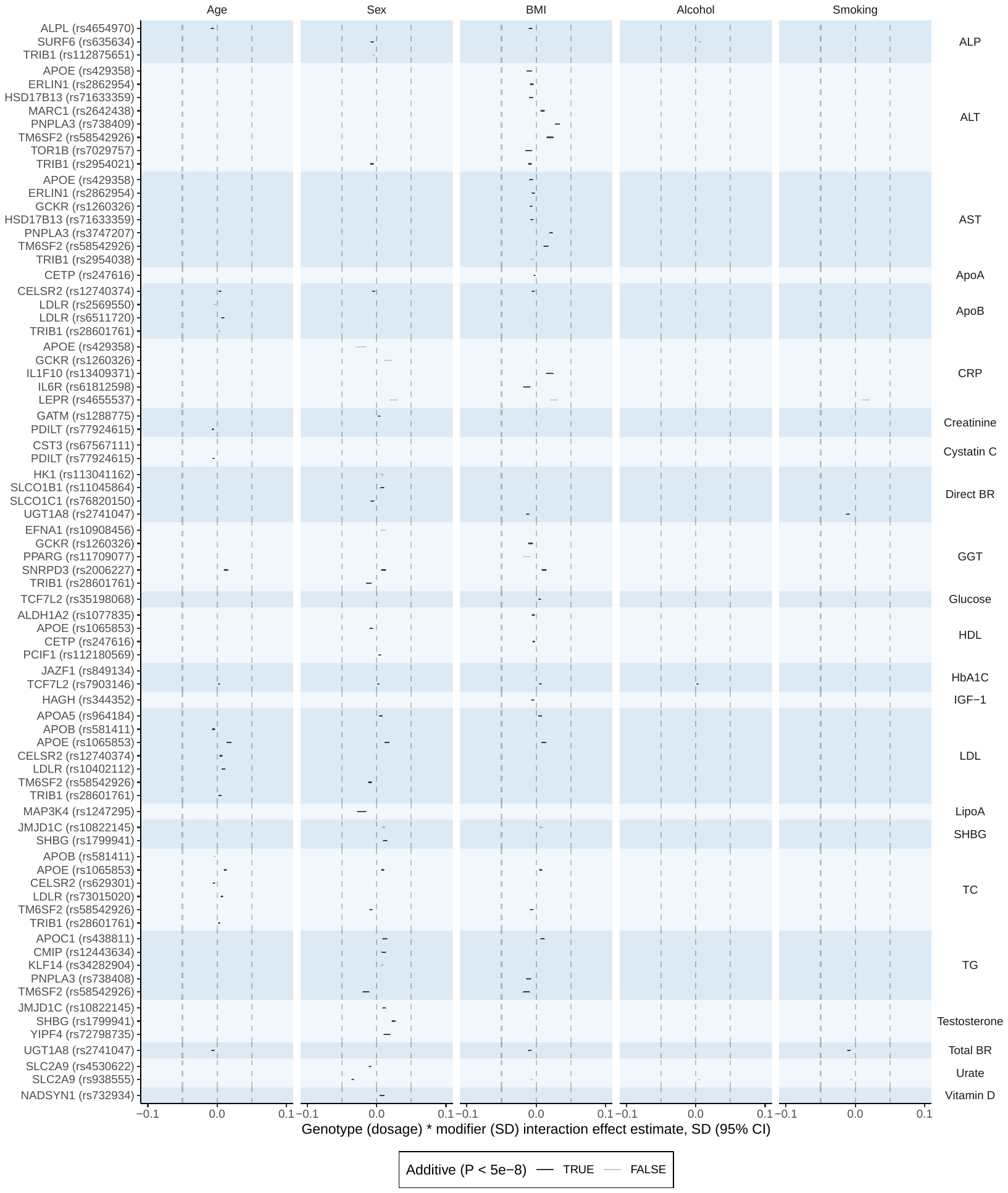

GxE effects using multiplicative scale and heteroscedasticity consistent standard errors^8^ (P < 5 x 10^-8^). ALP, alkaline phosphatase. ALT, alanine aminotransferase. AST, aspartate aminotransferase. ApoA, Apolipoprotein A. ApoB, apolipoprotein B. CRP, C-reactive protein. Direct BR, direct bilirubin. GGT, Gamma glutamyltransferase. HDL, high-density lipoprotein. HbA1c, glycated haemoglobin. LDL, low-density lipoprotein. LipoA, lipoprotein A. IGF-1, insulin-like growth factor 1. SHBG, sex-hormone binding globulin. TC, total cholesterol. TG, triglycerides. Total BR, total bilirubin. BMI, body mass index. Smoking, smoking status. Alcohol, intake. PA, physical activity. All measures reported on SD scale. All estimates were adjusted for the main effect, age, sex, and top ten genetic principal components. Gene name is the nearest protein coding gene HGNC name by chromosomal position. SD, standard deviation. CI, confidence interval.

**Figure S11. Top gene-by-environment interaction effects (P < 5 x 10^-8^) on biomarker concentration using additive scale adjusted for fine-mapped main effect**

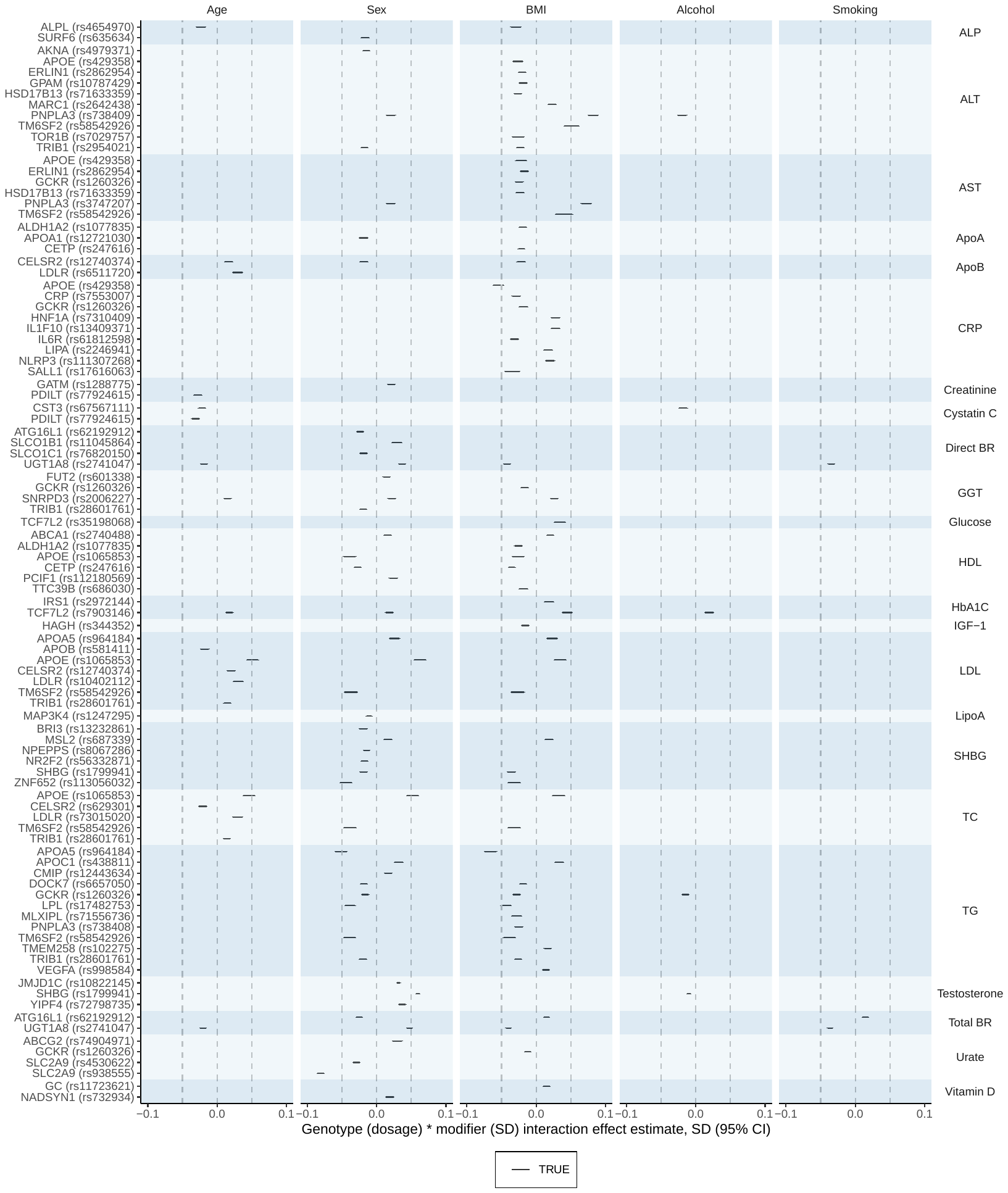

GxE effects using additive scale and heteroscedasticity consistent standard errors^8^ (P < 5 x 10^-8^) adjusted for fine-mapped main effects. ALP, alkaline phosphatase. ALT, alanine aminotransferase. AST, aspartate aminotransferase. ApoA, Apolipoprotein A. ApoB, apolipoprotein B. CRP, C-reactive protein. Direct BR, direct bilirubin. GGT, Gamma glutamyltransferase. HDL, high-density lipoprotein. HbA1c, glycated haemoglobin. LDL, low-density lipoprotein. LipoA, lipoprotein A. IGF-1, insulin-like growth factor 1. SHBG, sex-hormone binding globulin. TC, total cholesterol. TG, triglycerides. Total BR, total bilirubin. BMI, body mass index. Smoking, smoking status. Alcohol, intake. PA, physical activity. All measures reported on SD scale. All estimates were adjusted for the main effect, age, sex, and top ten genetic principal components. Gene name is the nearest protein coding gene HGNC name by chromosomal position. SD, standard deviation. CI, confidence interval.
**Figure S12. Top gene-by-gene interaction effects (P < 5 x 10^-8^) on biomarker concentration using additive scale**

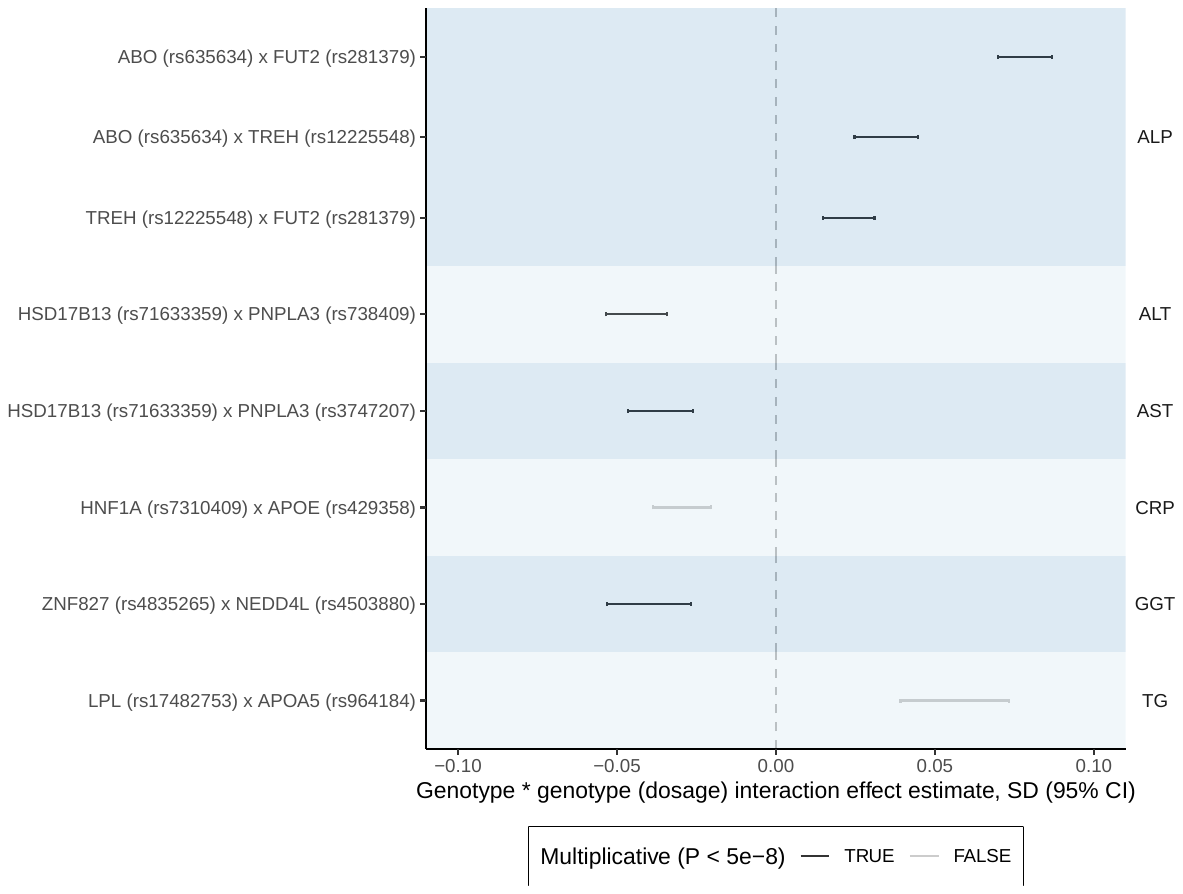

GxG effects using additive scale and heteroscedasticity consistent standard errors^8^ (P < 5 x 10^-8^) adjusted for the main effect, age, sex, and top ten genetic principal components. ALP, alkaline phosphatase. ALT, alanine aminotransferase. AST, Aspartate aminotransferase. CRP, C-reactive protein. GGT, Gamma glutamyltransferase. TG, triglycerides. All measures reported on SD scale. Gene name is the nearest protein coding gene HGNC name by chromosomal position. SD, standard deviation. CI, confidence interval.

**Figure S13. Top gene-by-gene interaction effects (P < 5 x 10^-8^) on biomarker concentration using multiplicative scale**

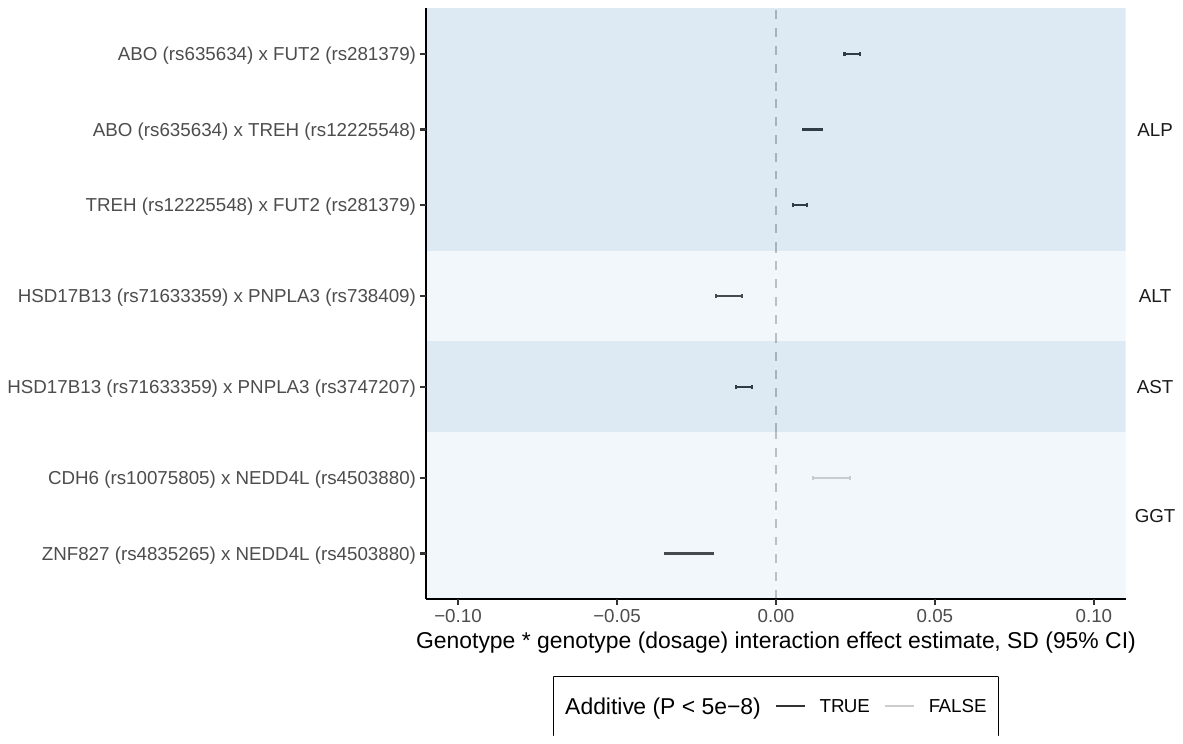

GxG effects using multiplicative scale and heteroscedasticity consistent standard errors^8^ (P < 5 x 10^-8^) adjusted for the main effect, age, sex, and top ten genetic principal components. ALP, alkaline phosphatase. ALT, alanine aminotransferase. AST, Aspartate aminotransferase. GGT, Gamma glutamyltransferase. All measures reported on SD scale. Gene name is the nearest protein coding gene HGNC name by chromosomal position. SD, standard deviation. CI, confidence interval.

**Figure S14. Top gene-by-gene interaction effects (P < 5 x 10^-8^) on biomarker concentration using additive scale adjusted for fine-mapped main effects**

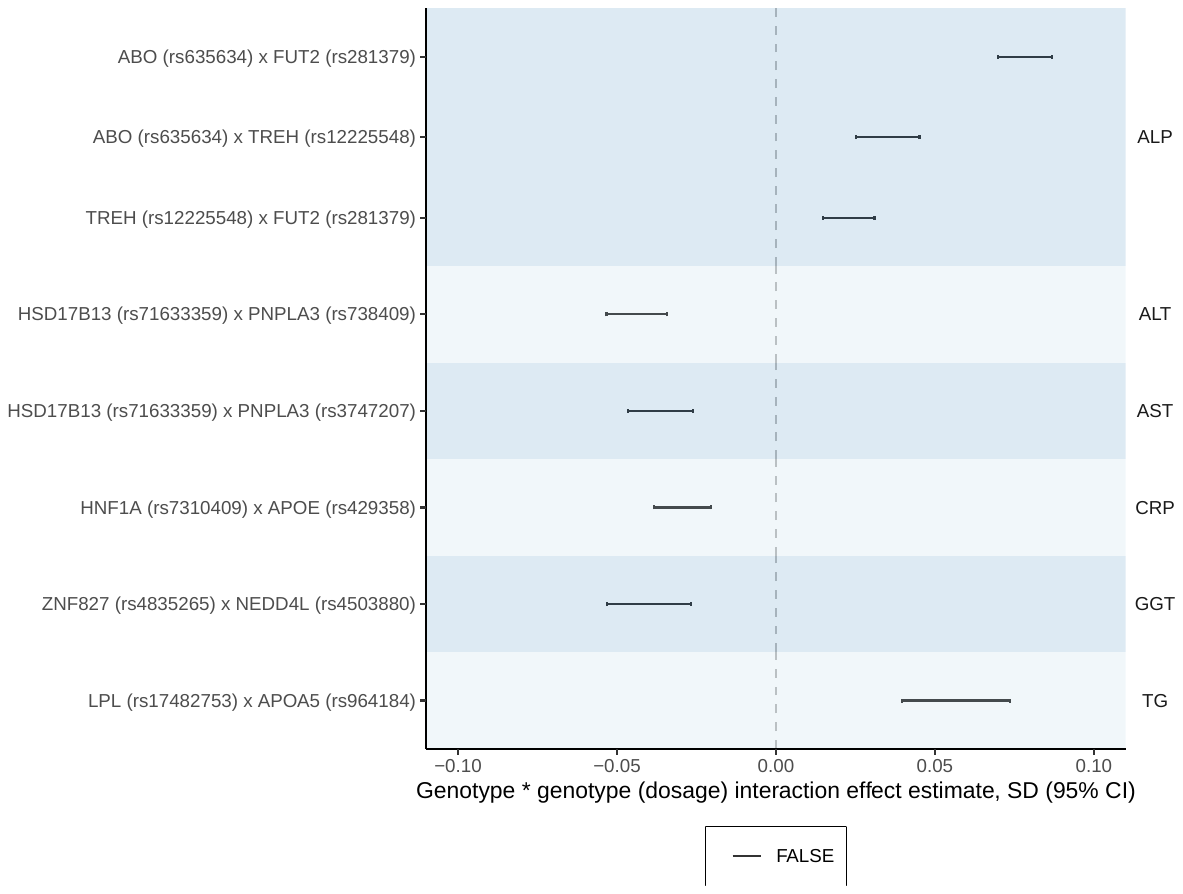

GxG effects using additive scale and heteroscedasticity consistent standard errors^8^ (P < 5 x 10^-8^) adjusted for the main effect, age, sex, top ten genetic principal components and fine-mapped main effects. ALP, alkaline phosphatase. ALT, alanine aminotransferase. AST, Aspartate aminotransferase. CRP, C-reactive protein. GGT, Gamma glutamyltransferase. TG, triglycerides. All measures reported on SD scale. Gene name is the nearest protein coding gene HGNC name by chromosomal position. SD, standard deviation. CI, confidence interval. **Table S1. GWAS summary statistics for top vQTLs identified through this study.** Clumped per allele SNP effects on biomarker (SD) variance adjusted for age, sex, and top ten genetic principal components in both models. Loci were identified using the regression-based Brown-Forsythe test (implemented in varGWAS) and experiment wise threshold (P < 5 x 10^-8^ / 30) on the additive scale. SNP, variant using genomic location based on GRCh37/hg19. Rsid, dbSNP identifier. Outcome, biomarker. Gene, nearest gene. Phi_x1, effect of one allele increase on biomarker variance. Se_x1, standard error for phi_x1. Phi_x2, effect of two allele increase on biomarker variance. Se_x2, standard error for phi_x2. Phi_f, F-statistic. Phi_p, P-value.

**Table S2. Top GxG/GxE effect summary statistics.** Per allele SNP interaction effect on biomarker concentration adjusted for age, sex, top ten genetic principal components using heteroscedastic consistent standard errors. SNP, nearest gene and dbSNP identifier. Outcome, biomarker. Beta, interaction effect size (SD). Lci, 95% confidence interval, lower interval. Uci, 95% confidence interval, upper interval. P, P-value for the interaction test.

**Table S3. Fine-mapped loci covariates.** List of fine-mapped SNPs used as covariates in gene-gene and gene-environment interaction sensitivity analyses. SNP, nearest gene and dbSNP identifier. Outcome, biomarker. Fine_mapped, pipe separated list of fine-mapped SNPs used as covariates. Each SNP has the from chromosome, position using GRCh37/hg19, non-effect allele, effect allele.
